# The minor spliceosome is a master immune regulator

**DOI:** 10.1101/2025.09.12.25335567

**Authors:** MB Johnson, J Russ-Silsby, PA Blair, M Govier, G Bonfield, C Domingo-Vila, EXE-T1D consortium, ATAC clinical consortium, MN Wakeling, RA Oram, SE Flanagan, TIM Tree, KA Patel, AT Hattersley, E De Franco

## Abstract

Pathogenic variants in non-coding genes are emerging as critical contributors to human rare diseases. We identified 19 individuals with early-onset diabetes (diagnosed <5 years) and additional clinical features who had biallelic pathogenic variants in the novel disease gene *RNU6ATAC* (n=7) or in *RNU4ATAC* (n=12). Both genes encode non-coding components of the minor spliceosome, a protein-RNA complex mediating splicing of ∼700 genes containing U12/minor-type introns. RNA-seq of whole blood from six patients showed aberrant splicing of minor intron-containing genes in individuals with *RNU6ATAC* (n=3) or *RNU4ATAC* (n=3) variants. 50% of patients tested were islet-autoantibody positive, and 12/19 had additional immune features of immune dysregulation. Analysis of patients’ transcriptomic, methylation and immune data revealed impaired B cell development and maturation. Biallelic disease-causing variants in *RNU6ATAC* and *RNU4ATAC* therefore cause syndromic early-onset autoimmune diabetes and immune dysregulation, highlighting the critical role of the minor spliceosome as a master regulator of the human immune system.

---

Uncovering novel non-coding causes of human disease provides key insights into the regulation of core biological processes in humans. Despite significant advances since the introduction of exome and genome sequencing, up to half of individuals affected by a rare disease remain without a genetic diagnosis (1,2). Variants in the non-protein coding genome—including regulatory elements such as promoters, enhancers, and untranslated regions—may account for a considerable fraction of this diagnostic gap (3–5). The recent discovery that ReNU syndrome, caused by variants in the non-protein-coding gene *RNU4-2*, explains ∼0.4% of cases with undiagnosed neurodevelopmental delay, highlighting the importance of these genes in rare disease (6–8).

Here, we report the identification of variants in the non-coding minor spliceosome component *RNU6ATAC* as a novel disease gene, causing early-onset autoimmune diabetes and hypogammaglobulinemia. We also extend the phenotype associated with biallelic variants in another non-coding minor spliceosome component, *RNU4ATAC,* to include early-onset autoimmune diabetes. Our results uncover the essential role of the minor spliceosome in regulating the human immune system.

## RESULTS AND DISCUSSION

To identify new genetic causes of autoimmune diabetes and immune dysregulation, we performed genome sequencing in three consanguineous individuals with infancy-onset diabetes (diagnosed aged <12 months) and hypogammaglobulinemia. All known genetic causes of infancy-onset diabetes had been previously excluded. No shared genes with ultra-rare homozygous coding variants (gnomAD v4.1.0 (9) MAF <1×10^-5^) in all 3 individuals were identified. We next looked for ultra-rare homozygous variants in non-coding genes. This identified a single common gene, the small nuclear RNA (snRNA) *RNU6ATAC* (Figure 1a). All three cases had a different *RNU6ATAC* homozygous variant that was absent from gnomAD v4.1.0 (∼75,000 individuals) and ClinVar (10). The affected sibling of one of the individuals (Family A) was also homozygous for the *RNU6ATAC* rare variant. *RNU6ATAC* has not been previously reported as a human monogenic disease gene.

**Figure 1:**
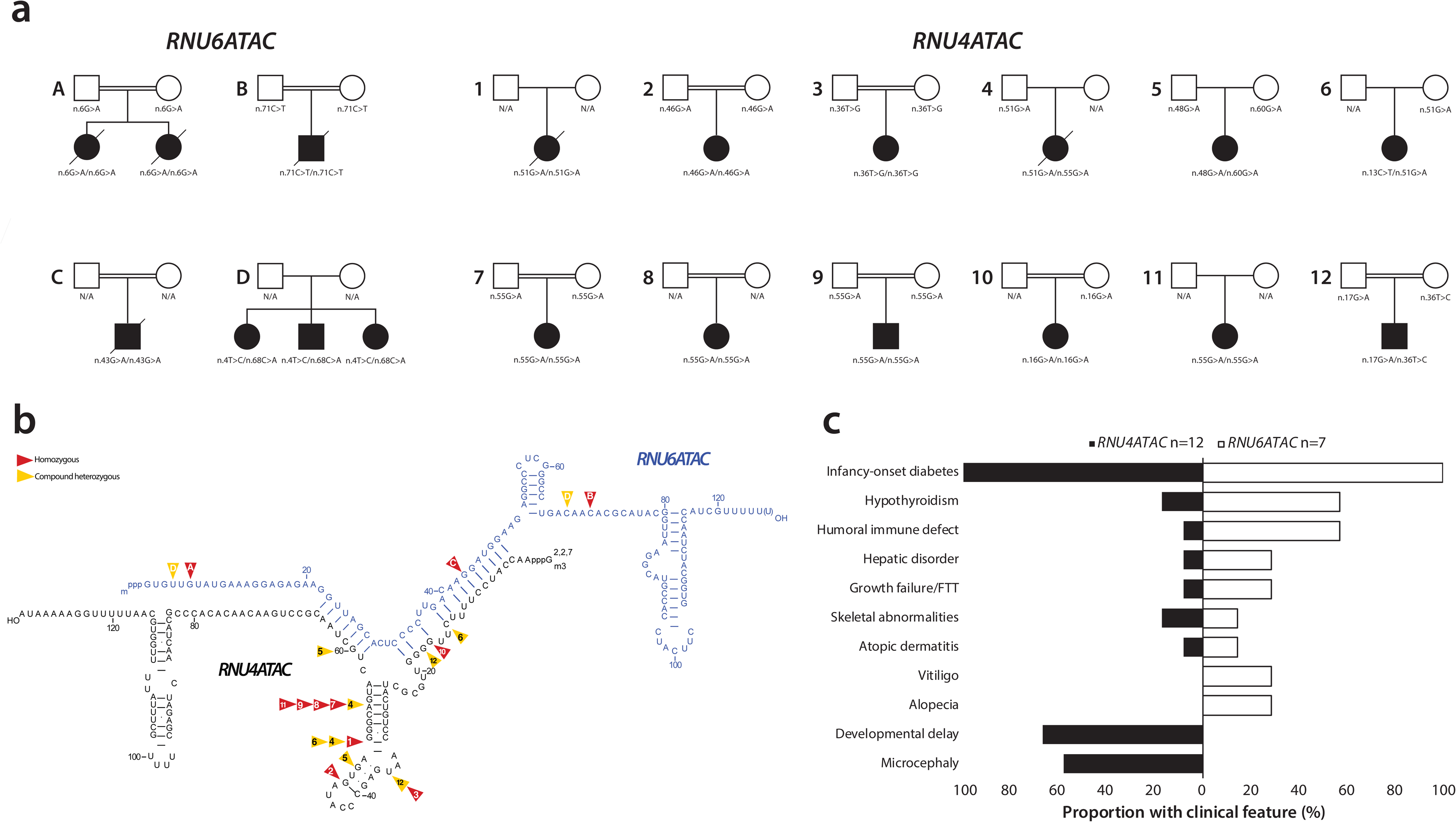
Genetic and clinical information for the RNU6ATAC and RNU4ATAC cohorts. A) Partial pedigrees of the cases with *RNU6ATAC* and *RNU4ATAC* biallelic variants. B) Secondary structure of RNU4ATAC and RNU6ATAC snRNA duplex with variant positions highlighted and number/letter by case. Heterozygous variants identified in trans with another pathogenic variant are shown with yellow arrows. Homozygous variants are shown with red arrows. Hydrogen bonds between bases in stem I and stem II are shown with lines. Adapted from (37). C) Tornado plot showing clinical features in *RNU6ATAC* vs. *RNU4ATAC* cohorts.

*RNU6ATAC* forms part of the catalytic core of the minor spliceosome (10,11). This fundamental and highly evolutionarily conserved protein-RNA complex mediates the excision of <0.5% of human introns containing the U12-type sequence motif to create mature mRNA (12). Disruption of the spliceosome causes intron retention that leads to downstream impacts including transcript loss due to nonsense mediated decay, or disruption to protein function through truncation or elongation (13).

We next investigated if biallelic variants in all 65 components of the minor spliceosome were implicated in the aetiology of early-onset diabetes (supplementary Table 1) (11,14). We screened for ultra-rare variants in these genes using existing genome sequencing data from 276 individuals with neonatal diabetes (diagnosed <6 months) or early-onset diabetes with additional autoimmune disease (both diagnosed <5y) in whom known causes of monogenic diabetes had been excluded (15). This identified 3 further individuals from 1 family with biallelic variants in *RNU6ATAC,* and 7 unrelated individuals with homozygous or compound heterozygous variants in *RNU4ATAC* (Figure 1a). We did not identify ultra-rare biallelic variants in any of the 63 remaining genes. We then Sanger-sequenced *RNU6ATAC* and *RNU4ATAC* in 196 patients with neonatal diabetes of unknown cause who had not undergone genome sequencing, this identified a further 5 cases with biallelic variants in *RNU4ATAC*.

*RNU4ATAC* binds directly to *RNU6ATAC* to stabilise the pre-catalytic configuration of the minor spliceosome, sequestering *RNU6ATAC’s* catalytic elements (10,11). Recessively inherited variants in *RNU4ATAC* are a known cause of ‘RNU4ATACopathies’ (16), encompassing a highly variable clinical spectrum that commonly includes microcephaly, developmental delay and intra-uterine growth restriction (IUGR), and can include immune dysregulatory features. Diabetes has not been reported as a feature of RNU4ATACopathies.

In total, we identified 19 individuals from 16 families with defects in the minor spliceosome and early-onset diabetes: 7 individuals from 4 families with biallelic *RNU6ATAC* variants and 12 unrelated individuals with biallelic *RNU4ATAC* variants (Figure 1a, Figure 1b). Variant testing confirmed carrier status in all the parents available for testing (n = 19).

### Clinical features

There was substantial overlap in clinical features between patients with variants in *RNU6ATAC* and *RNU4ATAC*, but also some notable differences (Fig. 1c). Biallelic *RNU6ATAC* variants caused early-onset diabetes (median onset: 17 weeks) with additional immune dysregulation in 6/7 cases. Common features included humoral immune defects (4/7; B-cell lymphopenia, a/hypogammaglobulinemia) and autoimmunity (4/7; hypothyroidism [n=4], alopecia [n=2]). Cholestatic jaundice and elevated liver enzymes were observed in two cases. Among cases with reported gestation, 3/5 had IUGR (Z-score < -1.28 (18)).

Individuals with biallelic *RNU4ATAC* variants often displayed classic RNU4ATACopathy features (16), including microcephaly, IUGR, and developmental delay, which were seen in 10/12 cases. Six had immune dysregulation (hypothyroidism [n=2], recurrent infections [n=3]). Autoimmunity (e.g., Addison’s disease, autoimmune hypothyroidism) has been reported in some RNU4ATACopthy cases (19), including a single individual with type 1 diabetes (T1D) and Addison’s disease (20). In our cohort, 12 unrelated individuals had early-onset diabetes (median onset: 20 weeks), confirming that diabetes is part of the RNU4ATACopathy spectrum. Diabetes is unlikely to be attributable to specific variants in *RNU4ATAC* as 12/15 variants identified in this study were previously reported to cause RNU4ATACopathies in patients who did not have diabetes (17).

Variants in *RNU6ATAC* and *RNU4ATAC* were associated with early-onset diabetes and immune dysregulation. Developmental abnormalities were common in *RNU4ATAC* cases but not *RNU6ATAC*, although later-onset developmental issues in the latter group cannot be excluded. A recent study identified a patient with compound heterozygous ultra-rare *RNU6ATAC* variants of unknown clinical significance (n.36T>G, n.28C>T) who had IUGR, postnatal growth failure, microcephaly, epilepsy, intellectual disability, and ataxia but no diabetes (21).

### RNA-seq showed U12 intron retention

To assess the impact of the identified *RNU6ATAC* and *RNU4ATAC* variants on U12-intron splicing, we obtained fresh whole-blood RNA samples from 6 cases: 3 unrelated individuals with *RNU4ATAC* variants and 3 affected siblings from one *RNU6ATAC* family. We performed RNA-seq on these samples, 9 unaffected parents (4 *RNU6ATAC*, 5 *RNU4ATAC*), 4 age-matched healthy and 4 age-matched T1D controls (Supplementary Table 2). This identified significant intron retention in both *RNU4ATAC* and *RNU6ATAC* cases compared to controls (Figure 2a). The majority of genes with significant intron retention (n=258/275, 94%) were known U12-intron-containing genes listed in the Intron Annotation and Orthology U12 database (IAOD) (Figure 2b, Supplementary Table 3) (22). The remaining 17 genes likely represent novel U12 genes given we found significant intron retention in individuals with biallelic variants in either *RNU4ATAC* (n=1), *RNU6ATAC* (n=4) or both (n=12) (Figure 2a, Supplementary Figure 1, Supplementary Table 3). The high level of intron retention of U12 genes was similar between individuals with *RNU6ATAC* and *RNU4ATAC* variants and comparable to that previously reported in RNU4ATACopthy cases (19,23,24).

**Figure 2:**
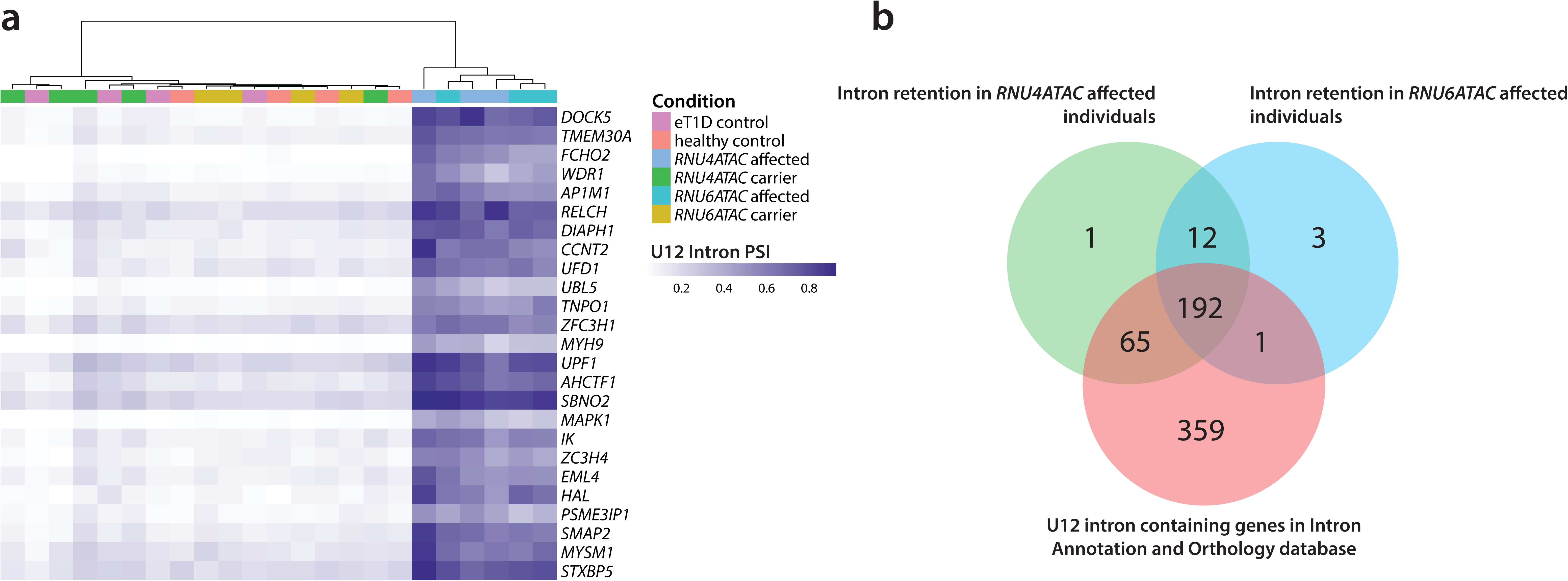
RNA-seq identifies significant intron retention in both RNU6ATAC and RNU4ATAC cases. A) Heatmap showing top 25 genes with significant intron retention. Both *RNU4ATAC* and *RNU6ATAC* cases cluster together, showing similar profiles of intron retention across U12-intron containing genes. B) Venn diagram showing the intersection of known U12-intron containing genes (IAOD (22)) (green), and empirically defined genes with significant intron retention in whole blood RNA from *RNU6ATAC* cases (n=3, pink) and *RNU4ATAC* cases (n=3, blue). Thirteen genes were identified which showed significant intron retention in both cohorts but were not present in the IAOD database, likely reflecting newly identified U12 genes.

### Clinical and biomarker data support an autoimmune diabetes aetiology

To investigate the diabetes mechanism, we first performed islet autoantibody testing in available serum samples (n=4 *RNU6ATAC*, n=6 *RNU4ATAC*). Five of the ten individuals (50%) were positive for GADA antibodies (threshold >97.5^th^ centile of the non-diabetic population). This rate of positive islet autoantibodies is similar to that seen in age-matched T1D and known monogenic autoimmune diabetes cases (25,26), indicating that the diabetes in these individuals is likely driven by islet autoimmunity. Furthermore, in cases with available clinical data, all individuals presented in infancy/early-childhood with very high glucose values (median 32mmol/L, IQR 25-39, n=15) and were insulin treated with full replacement doses (median 1.0 U/Kg/day, IQR 0.7-1.5, n=13), indicative of profound and rapid loss of endogenous insulin secretion.

### Multi-omic analysis of patient samples revealed a profound B cell defect

To investigate potential mechanisms for the autoimmune diabetes and additional immune dysregulatory features, we performed Weighted Gene Co-expression Network Analysis on our RNA-seq data (27). This identified 2 significantly differentially regulated gene modules in *RNU4ATAC*/*RNU6ATAC* cases compared to controls (Figure 3a). Enrichment analysis through Gene Ontology (28,29) and KEGG (30) converged on significant enrichment for genes involved in B cell signalling, development or proliferation and innate immune responses, as well as other immune pathways (Figure 3b, Supplementary Figure 2).

**Figure 3:**
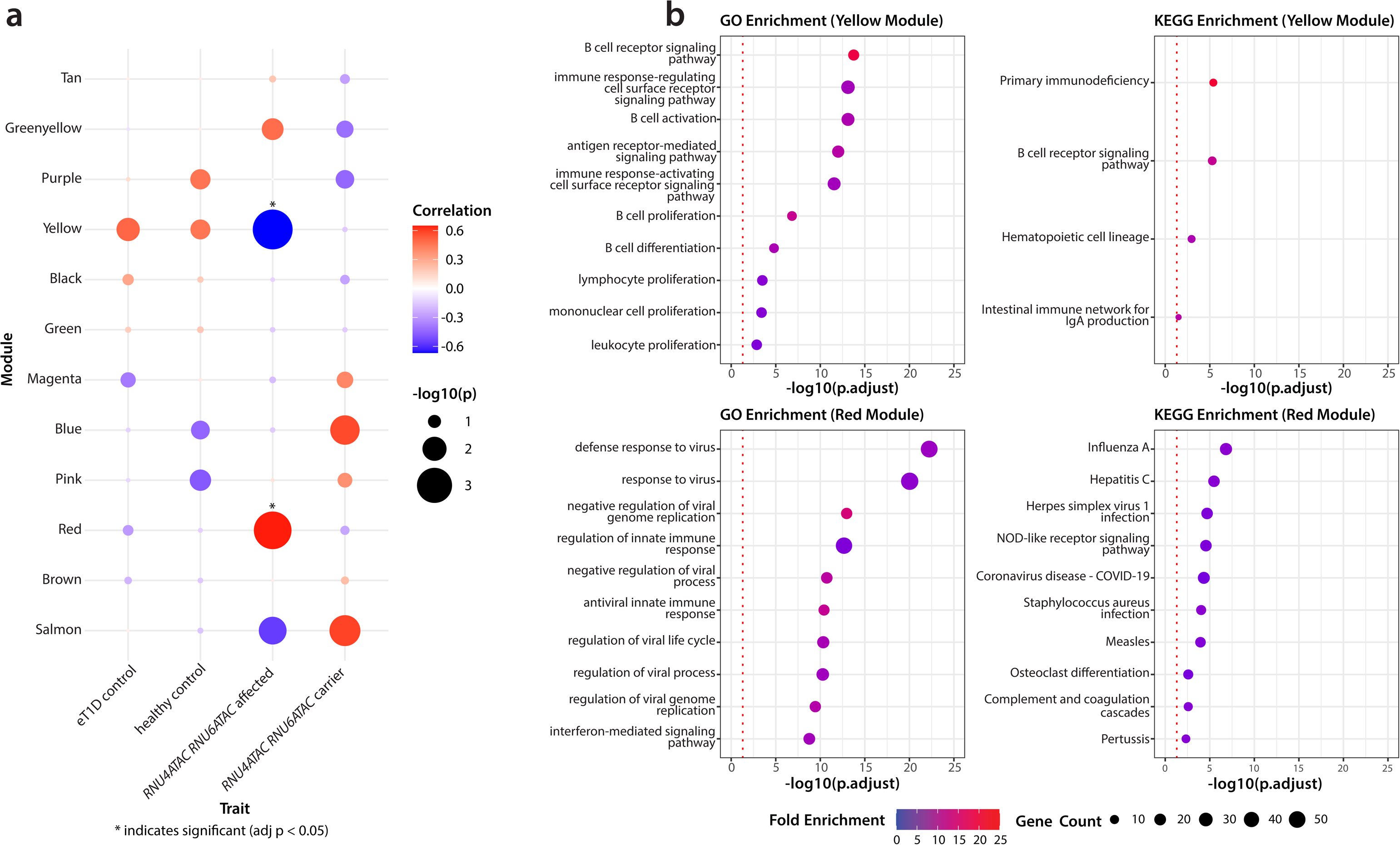
Weighted Gene Co-Expression Network Analysis and gene enrichment analysis of resulting enriched gene modules from patient RNA-seq data. A) Correlation between weighted gene co-expression network analysis (WGCNA) and derived gene modules and disease status, which included affected individuals with *RNU6ATAC* and *RNU4ATAC* variants, carriers, age matched T1D controls and age matched healthy controls. Module names are arbitrarily defined. Significant correlation after Bonferroni correction for multiple testing shown by *. B) GO and KEGG enrichment analysis of significantly correlated gene modules with log transformed adjusted P value shown. Enriched pathways converge on immune defects, particularly B cell and other humoral immunity.

To investigate the impact on B cells we performed deconvolution analysis of our whole blood genome-wide methylation data (31). This showed significantly reduced estimated naïve B cells (Figure 4a), whilst memory B cells and 10 other immune cell subsets did not show significant differences to controls (Supplementary Figure 3). To validate these findings, we performed in-depth immune profiling on an individual with biallelic *RNU4ATAC* variants (case 6, Figure 1a) in whom we were able to obtain a fresh whole blood sample. This showed a striking B cell developmental defect, with reduced naïve and memory B cells and increased transitional B cells and antibody secreting cells vs. age matched healthy and T1D controls (figure 4b). This suggests impaired B cell development and maturation. We also found reduced basophils and increased activation of CD8+ and CD4+ T cells, while other cell types were similar to healthy and T1D controls (Supplementary Figure 4). The patient was not known to have an infection when the sample was taken, but we are unable to rule out a nascent infection that could explain these findings.

**Figure 4:**
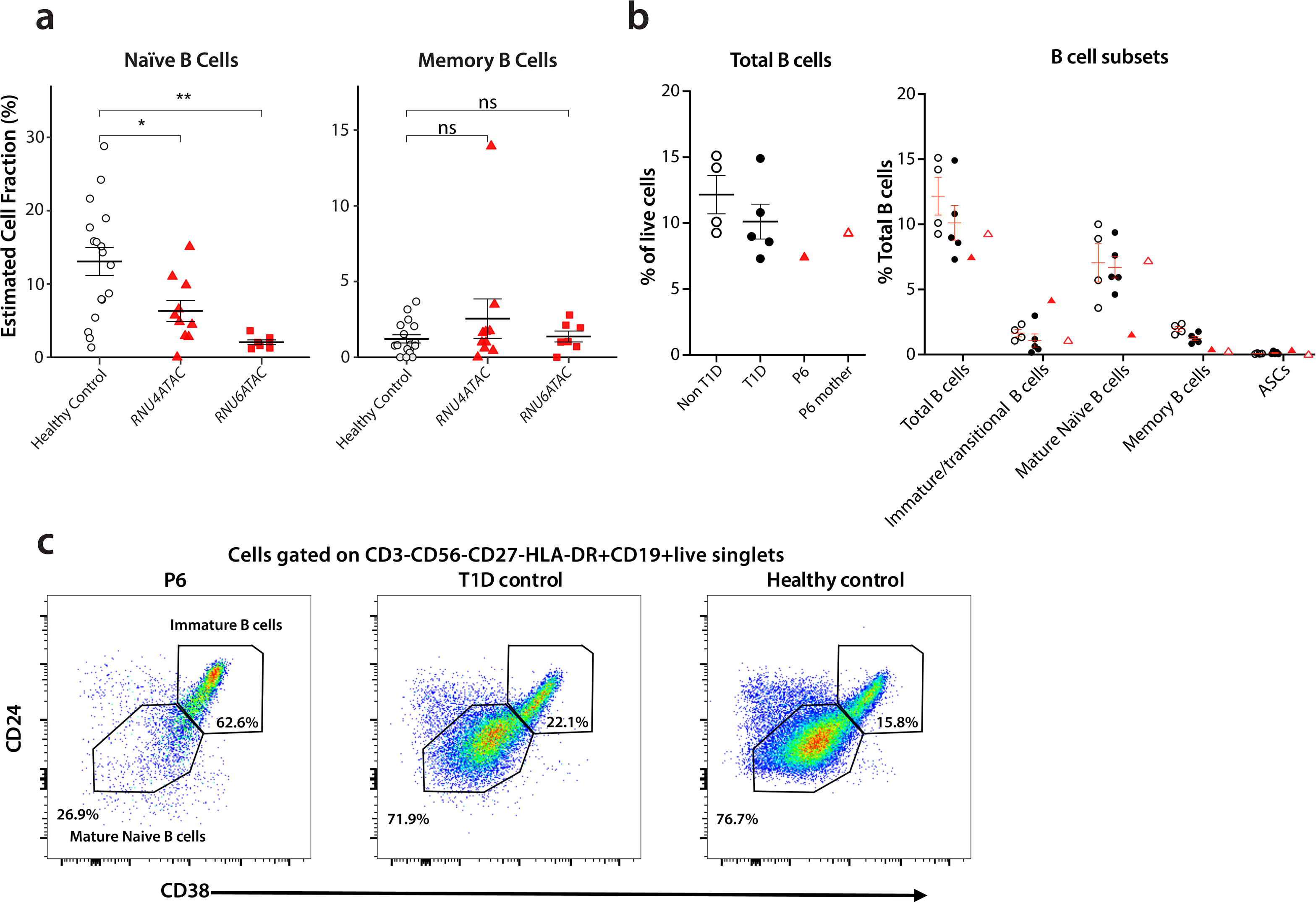
Immune profile analysis of patients using methylation and flow cytometry. A) Deconvolution of whole-blood derived DNA suing the IDOL library identified normal estimates of memory B cells but significantly reduced naïve B cells in both RNU4ATAC (n=10) and RNU6ATAC (n=7) cases compared to age matched controls. B) Flow cytometry on fresh blood cells of an *RNU4ATAC* patient (red triangle) identified reduced naïve and increased transitional B cells, as well as elevated antigen presenting (ASC) cells, compared to healthy controls (white circles) and age matched T1D (black circles). C) Dot plots of flow cytometry data showing markedly increased immature B cell compartment and reduced mature naive B cells in the RNU4ATAC case vs. representative controls.

## DISCUSSION

We demonstrate that biallelic pathogenic variants in *RNU6ATAC* cause a novel genetic syndrome characterised by monogenic autoimmune diabetes with additional immune dysregulation. We also extend the phenotype associated with biallelic variants in *RNU4ATAC* to include early-onset autoimmune diabetes. These represent the first causes of monogenic diabetes resulting from pathogenic variants in non-protein-coding genes.

Our results show a key role for the minor spliceosome in immune regulation. We identified 19 individuals with defects in minor spliceosome components; 7 with the novel *RNU6ATAC* associated syndrome and 12 with *RNU4ATAC* variants (n=12), all of whom have early-onset diabetes and 12 (63%) of whom have additional immune dysregulatory features. Using a multi-omic approach combined with biomarkers, we have triangulated evidence that the diabetes in these individuals is autoimmune and identified a shared B cell developmental defect across both monogenic disorders.

Our data highlights profound B cell defects resulting from pathogenic variants in *RNU4ATAC* or *RNU6ATAC*. This is consistent with B cell defects previously reported in some patients with RNU4ATACopathies without diabetes, particularly in the Roifman syndrome subtype (16). The role of B cells in the pathogenesis of islet autoimmunity is debated; some evidence points to a direct pathogenic role, whilst other evidence suggests B cell dysregulation is secondary to islet autoimmunity (32). Postmortem pancreas of individuals with the young onset ‘TIDE1’ endotype of T1D show increased insulitic B cells (33), and B cell depletion with the monoclonal antibody Rituximab has shown some success in delaying T1D progression (34). However, a patient with absent B-cells due to X-linked agammaglobulinemia (XLA), developed autoimmune diabetes implying that B cells or autoantibodies are not required for diabetes development (35). Further study of patients with both B cell defects and autoimmune diabetes is warranted to understand the role of B cell regulation, maturation and development in autoimmune diabetes.

The minor spliceosome is found across many eukaryotes, though has been lost in some genera (12). Whilst it is involved in exon excision for a minority of genes/introns, many have essential functions and minor intron splicing may play a role in temporal regulation of protein expression through slower mRNA processing (36). This is supported by incomplete intron retention of U12 genes as seen in our cases and previously in RNU4ATACopathy (23,37). Further work is needed to untangle the role of genes undergoing minor splicing in the development of beta cell autoimmunity, as it is possible that a subset of these genes have direct pathogenic roles.

We performed methylation analysis on 17 cases (7 *RNU6ATAC* and 10 *RNU4ATAC*) and RNA-Seq on 6 cases (3 and 3). The results of these studies supported all individuals having B cell defects which were further validated in a single *RNU4ATAC* case using gold-standard immune analyses through flow cytometry. Although we would have liked to study more individuals, the geographic diversity of the cohort and the severity of disease (6 cases deceased in early life) prevented sample collection from the remaining patients.

In conclusion, we have identified 19 individuals with biallelic pathogenic variants in the non-coding minor spliceosome genes *RNU6ATAC* and *RNU4ATAC*, resulting in syndromic early-onset autoimmune diabetes. This work provides key insights into the role of the minor spliceosome in human immune regulation, new mechanistic insights into pathways resulting in beta-cell autoimmunity and crucial new diagnoses to families.

## METHODS

### Subjects

The study was conducted in accordance with the Declaration of Helsinki and all subjects, or their parents, gave informed consent for DNA extraction, genetic testing and sample storage in the Genetic Beta Cell Research Bank (https://www.diabetesgenes.org/current-research/genetic-beta-cell-research-bank/). The study was approved by the Wales Research Ethic Committee 5 Bangor (REC 17/WA/0327, IRAS project ID 231760). Individuals with neonatal diabetes (NDM: diagnosed <6 months) or early-onset diabetes (diagnosed <5 years) were recruited by their clinicians to the Exeter Genomics Laboratory for monogenic diabetes genetic testing through a dedicated referral form (https://www.diabetesgenes.org/download/3564/?tmstv=1715331423). Genetic ancestry was assigned using Procrustes analysis and random forest classification (38).

### Genetic testing

Whole-genome sequencing of DNA extracted from peripheral blood leukocytes was completed on 181 individuals with NDM and 95 with early-onset diabetes (n=70 with Illumina HiSeq X10 [illumina, USA] n=206 with BGISeq-500 [BGI Europe, Poland]). The resulting sequence reads were aligned to GENCODE release 48 (GRCh38.p14) using BWA MEM version 0.7.15 (39,40), and processed with our bespoke pipeline based on GATK best practices (Picard version 2.7.1 and GATK version 3.7 (41)). All samples had mean coverage >30x and >95% coverage at >20x. Variants were annotated using Alamut batch standalone version 1.11 (SOPHiA Genetics, Switzerland). We separately analysed the 19,435 coding and 59,251 non-coding genes as annotated in the GENCODE reference (42). For replication and family member testing we performed direct Sanger sequencing of *RNU4ATAC* and *RNU6ATAC* following PCR using the following primers:

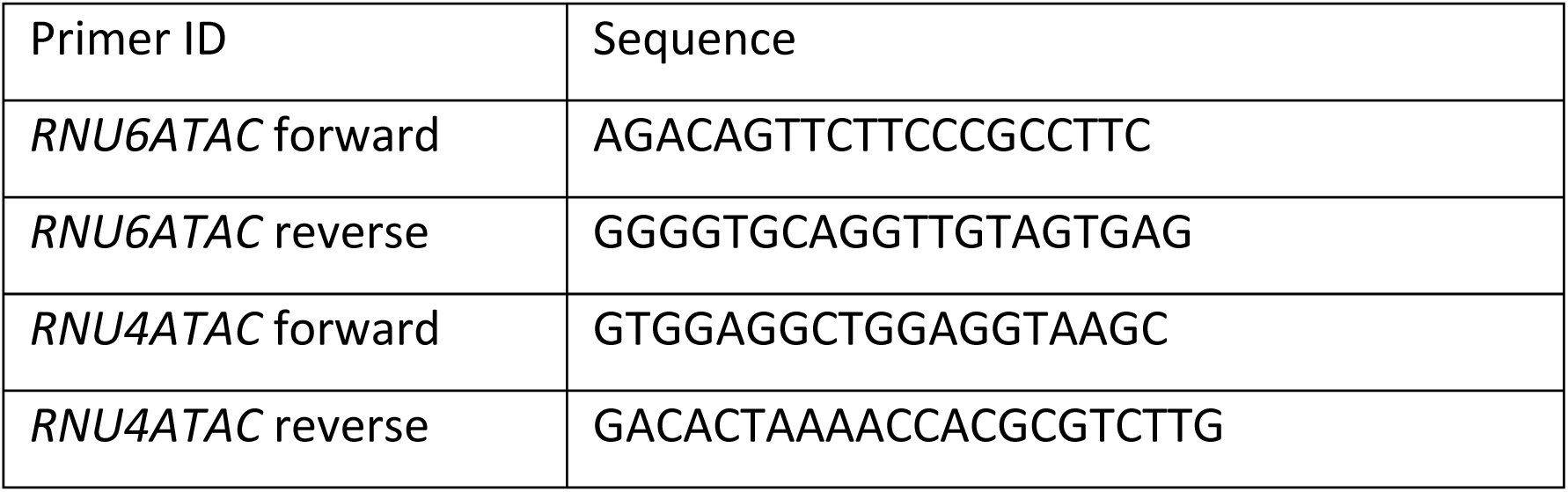

### Islet autoantibody testing

Serum Islet autoantibody testing (GADA, IA-2A, ZnT8A) was performed by Enzyme linked immunosorbent assay (ELISA), at Exeter Clinical Laboratory International (https://www.exeterlaboratory.com/blood-sciences/). This laboratory is UKAS accredited (ISO 15189:2012) and participates in UKNEQAS accreditation and the Islet Autoantibody Standardisation Program.

### Transcriptomics

Whole blood RNA was extracted from patient samples preserved in Tempus solution (Applied Biosystems, USA). RNA-seq experiments were performed at the Exeter Sequencing Facility (University of Exeter, UK). Briefly, 100ng RNA was prepared using the Illumina RiboZero library preparation kit to manufacturer’s instructions, followed by paired-end 100bp sequencing on an Illumina NovaSeq (Illumina, USA). The resulting reads were then aligned to the GRCh38 reference using STAR v2.7.11b (43). Transcripts were quantified using RSEM v1.33 (44) and intron retention was measured using SpliceWiz v1.10.1 (45). Weighted gene co-expression analysis was performed using the WGCNA package in R (46) to identify gene modules that were significantly correlated with affected status for minor spliceosomeopathies. Gene ontology (28,29) and KEGG pathway analysis (30) was then performed on the gene sets for these modules using the clusterProfiler package. Differential splicing analysis was performed on the intron retention values from SpliceWiz using EdgeR v4.6.2 (47).

### Methylation analysis

To investigate the original immune cell components of patient whole blood DNA samples we performed methylation array analysis using the EPIC v2 array on an iScan instrument (Illumina, USA). Initial QC was performed to ensure median methylated and unmethylated signal intensities were greater than 1000, that median bisulfite conversion percentage was above 80 and that sample sex determined from methylation data matched reported sex. Normalisation was performed using the Subset-quantile Within Array Normalisation (SWAN) method with the Minfi package (48). Finally, blood cell deconvolution was performed using the deconvolution tool and optimized blood cell methylation reference libraries from Salas et al. (31). This method estimates the proportion of cells in the original whole blood from 12 immune cell subsets (neutrophils, eosinophils, basophils, monocytes, naïve and memory B cells, naïve and memory CD4+ and CD8+ T cells, natural killer, and T regulatory cells) using measurement of methylation of cell-type specific loci.

### Flow cytometry

Leukocyte populations were characterised from fresh whole blood (collected in 2.7 mL EDTA tubes) using five multi-parameter flow cytometry panels previously validated and assessed for technical reproducibility across multiple laboratories (51–53). In brief, surface marker staining (Supplementary Table 5) of 100-200μL well-mixed fresh whole blood was performed for 45 min at RT, followed by red blood cell lysis during 8 min at RT (10x BD FACS lysing solution diluted in ddH2O, BD Biosciences, US). Staining for the lineage flow cytometry panel was conducted on a BD Trucount tube (BD Biosciences, US) enabling the calculation of absolute cell numbers alongside cell frequencies. The lineage tube was vortexed to ensure homogenisation and left on ice until acquisition (lyse, no wash). All other staining panel tubes were centrifuged at 500g for 5 min, cells were washed in 2 mL of FACS buffer [1x PBS (Invitrogen, US) containing 0.2% BSA (Sigma-Aldrich, US) and 2 mM EDTA (Sigma-Aldrich, US)] by centrifugation at 500g for 5 min, resuspended in 200 μL of FACS buffer and kept on ice until acquisition. Intracellular marker staining of 100 μL of fresh whole blood consisted of two 15 min incubations at RT with CD45RA-BV785 and 10 μL of fixative reagent (buffer 1 from PerFix-nc kit, Beckman Coulter, USA), respectively. Permeabilising reagent (buffer 2 from PerFix-nc kit, Beckman Coulter, USA) was added to the intracellular master mix of fluorescently labelled antibodies prepared previously, and cells stained intracellularly for 60 min at RT. After incubation, 3 mL of plain 1x PBS (Invitrogen, USA) was added for 5 min followed by centrifugation at 500g for 5 min. Cells were washed in 3 mL of 1x R3 reagent (10x buffer 3 diluted in ddH2O from PerFix-nc kit, Beckman Coulter, US), resuspended in 200 μL of 1x R3 reagent and kept on ice until acquisition. Phenotypes used to define cell subtypes are provided in supplementary table 6. The resulting flow cytometry data was analysed using FlowJo (BD Biosciences, USA) and compared to data from 4 age matched healthy controls and 5 age-matched T1D controls (supplementary table 7).

## Data availability

Anonymised methylation and RNA-seq data will be made available at the European Genome-phenome Archive web portal upon submission (https://ega-archive.org). Access to these data will be granted for appropriate use in research and will be governed by the provisions laid out in the terms contained in the Data Access Agreement. All other non-clinical data analysed during this study are included in this published article and supplementary information files. Clinical and genotype data can be used to identify individuals and is therefore available through collaboration to experienced teams working on approved studies examining the mechanisms, cause, diagnosis and treatment of diabetes and other beta-cell disorders. Requests for collaboration will be considered by a steering committee following an application to the Genetic Beta Cell Research Bank (https://www.diabetesgenes.org/current-research/genetic-beta-cell-research-bank/). Contact by email should be directed to Prof Elisa De Franco. All requests for access to data will be responded to within 14 days. Code used for analysis of methylation and RNA-seq data is available on GitHub (https://github.com/JamesR-S/RNU6ATAC_RNU4ATAC_Monogenic_Diabetes).

## Acknowledgements

We are grateful to the patients and their families for taking part in our gene discovery study. We thank Sabrina Wright and the Exeter Sequencing Facility (University of Exeter) for technical assistance, and Joe Burrage and Dr Emma Dempster for generating DNA methylation data. We are grateful to Dr Partick Willems and the GENDIA (Antwerp, Belgium) team for patient referral and for providing clinical details. This study was supported by the National Institute for Health and Care Research Exeter Biomedical Research Centre and the National Institute for Health and Care Research Exeter Clinical Research Facility. The views expressed are those of the authors and not necessarily those of the National Institute for Health and Care Research or the Department of Health and Social Care.

## Author information

These authors contributed equally: Matthew B Johnson, James Russ-Silsby.

## Contributions

M.B.J, J.R-S, A.T.H. and E.D-F designed the study, analysed and interpreted the data, wrote the manuscript and directed the project. P.A.B., C.D-V and T.I.M.T. performed immunological experiments, interpreted the resulting data and wrote the manuscript. M.G. and G.B. performed experiments and interpreted the resulting data. M.N.W and J.R-S wrote scripts to analyse genome sequencing data. M.B.J., S.E.F., K.A.P., E.D-F and A.T.H. and the ATAC-clinical consortium recruited patients and interpreted clinical data. M.J. and E.D.F. interpreted sequence variants. R.A.O. interpreted clinical data and recruited patients for flow. The EXE-T1D consortium contributed to recruitment of cases and controls for immune studies. All authors contributed to drafting the final manuscript. E.D.F. is the guarantor of the study and data.

