## Supplementary material for "The minor spliceosome is a master immune regulator"

1. EXE-T1D consortium members
2. ATAC clinician consortium members
3. Supplementary tables
    1 Minor spliceosome component genes
    2 Sample details for RNA-seq analyses
    3 U12 intron-containing genes
    4 Sample details for methylation analyses
    5 Antibodies used in flow cytometry analysis
    6 Markers used to define cell types from flow cytometry analysis
    7 Controls used in flow cytometry analysis
4. Supplementary figures
    1 Intron retention of new U12 genes
    2 WGCNA analysis of RNAseq data
    3 Estimates of immune cell proportions from deconvolution of methylation data
    4 Immune phenotyping of *RNU4ATAC* case

### 1. EXE-T1D CONSORTIUM

R A Dobbs[1], E Williams[2], K M Gillespie[3], W A Hagopian[4,5], A M Luckett[1], M Hudson[1], T J McDonald[1], N G Morgan[1], K Murrall[1], S Ramchand[1], S J Richardson[1], B O Roep[6], B Dimos[7], M E Smithmyer[7], C Speake[7]

1. Department of Clinical & Biomedical Sciences, University of Exeter Medical School, Exeter, UK
2. Department of Immunobiology, School of Immunology & Microbial Sciences (SIMS), King's College, London, UK
3. Translational Health Sciences, Bristol Medical School, University of Bristol, Southmead Hospital, Bristol, UK
4. Department of Paediatrics, Indiana University School of Medicine, Indianapolis, IN, USA.
5. Department of Medicine, University of Washington, Seattle, WA, USA.
6. Department of Internal Medicine, Leids Universitair Medisch Centrum, Leiden, Netherlands
7. Center for Interventional Immunology, Benaroya Research Institute, Seattle, WA

### 2. ATAC CLINICAL CONSORTIA

E Fink-Leinweber[1], M Lundgren[2,3], A Carlsson[4], M Al Mahdi[5], G Fadiana[6], F Soesanti[6], E A Mann[7], M T Bekx[8], T Randell[9], T Kontbay Çetin[10], M M Amoli[11], Can Thi Bich Ngoc[12], D C Vu[12], N H Lan[12], S S Albayati [13], N Thuse[14], K Jog[14], C Yajnik[14], K N Humayun[15], P Willems[16], A Djermane[17, 18], Y Ouarezki[17, 18]

1 - Märkische Kliniken GmbH Klinikum Lüdenscheid, Lüdenscheid, Germany
2 - Department of clinical sciences Malmö, Lund university, Sweden
3 - Skåne university Hospital, Kristianstad, Sweden
4 - Department of Pediatrics, Skånes University Hospital, Lund University, Sweden
5 – Dasman Diabetes Institute, Kuwait City, Kuwait
6 - Child Health Department, Faculty of Medicine, Universitas Indonesia, Cipto Mangunkusumo General Hospital, Jakarta, Indonesia
7 - Division of Endocrinology and Diabetes, Department of Pediatrics, University Hospital, Madison, US
8 - American Family Children's Hospital, University of Wisconsin-Madison, Madison Wisconsin
9 - Nottingham University Hospitals NHS Trust, Nottingham, UK
10 - Sanliurfa research and training hospital, Sanliurfa, Turkey
11 - Metabolic Disorders Research Centre, Endocrinology and Metabolism Molecular-Cellular Sciences Institute, Tehran University of Medical Sciences, Tehran, Iran
12 - Center for Endocrinology, Metabolism, Genetics/Genomics and Molecular Therapy, National Children's Hospital, Hanoi, Vietnam
13 - Child central teaching hospital, Baghdad, Iraq
14 - Diabetes Unit, KEM Hospital and Research Centre, Pune, India
15 - Aga Khan University, Karachi, Pakistan
16 - Gendia, Antwerp, Belgium
17 - EPH Hassan Badi, El-Harrach, Algiers, Algeria
18 - Université de Sciences de la Santé, Faculté de médecine d'Alger, Algeria

### 3. SUPPLEMENTARY TABLES

### **Supplementary table 1:** minor spliceosome genes. Taken from [ref]

| Protein | Subcomplex | Gene | Chr | Start | End | OMIM Inheritance | OMIM disease(s) |
|---|---|---|---|---|---|---|---|
| PRP8 | U5 snRNP | *PRPF8* | 17 | 1553923 | 1588176 | Autosomal Dominant | RETINITIS PIGMENTOSA 13 |
| SNU114 | U5 snRNP | *EFTUD2* | 17 | 42927311 | 42977030 | Autosomal Dominant | MANDIBULOFACIAL DYSOSTOSIS, GUION-ALMEIDA TYPE |
| BRR2 | U5 snRNP | *SNRNP200* | 2 | 96940074 | 96971297 | Autosomal Dominant | RETINITIS PIGMENTOSA 33 |
| U5-40K | U5 snRNP | *SNRNP40* | 1 | 31732417 | 31769662 | - | None |
| PRP6 | U5 snRNP | *PRPF6* | 20 | 62612488 | 62664453 | Autosomal Dominant | RETINITIS PIGMENTOSA 60 |
| DIM2 | U5 snRNP | *TXNL4B* | 16 | 72078188 | 72128330 | - | None |
| PRP28 | U5 snRNP | *DDX23* | 12 | 49223547 | 49246625 | - | None |
| SmD3 | U5 snRNP,U11 snRNP, U12 snRNP & U4atac/U6atac di-snRNP | *SNRPD3* | 22 | 24951471 | 25005947 | - | None |
| SmB | U5 snRNP,U11 snRNP, U12 snRNP & U4atac/U6atac di-snRNP | *SNRPB* | 20 | 2442280 | 2451499 | Autosomal Dominant | CEREBROCOSTOMANDIBULAR SYNDROME |
| SmD1 | U5 snRNP,U11 snRNP, U12 snRNP & U4atac/U6atac di-snRNP | *SNRPD1* | 18 | 19192228 | 19210417 | - | None |
| SmD2 | U5 snRNP,U11 snRNP, U12 snRNP & U4atac/U6atac di-snRNP | *SNRPD2* | 19 | 46190712 | 46195827 | - | None |
| SmF | U5 snRNP,U11 snRNP, U12 snRNP & U4atac/U6atac di-snRNP | *SNRPF* | 12 | 96252706 | 96297606 | - | None |
| SmE | U5 snRNP,U11 snRNP, U12 snRNP & U4atac/U6atac di-snRNP | *SNRPE* | 1 | 20383073<br>1 | 203839678 | Autosomal Dominant | Hypotrichosis 11 and Microcephaly(pending confirmation) |
| SmG | U5 snRNP,U11 snRNP, U12 snRNP & U4atac/U6atac di-snRNP | *SNRPG* | 2 | 70508494 | 70520903 | - | None |
| SCNM1 | U12 snRNP | *SCNM1* | 1 | 15112914<br>0 | 151142773 | Autosomal Recessive | OROFACIODIGITAL SYNDROME XIX |
| SF3b155 | U12 snRNP | *SF3B1* | 2 | 19825450<br>8 | 198299815 | Somatic | myelodysplastic syndrome(somatic mutations) |
| SF3b145 | U12 snRNP | *SF3B2* | 11 | 65818200 | 65836779 | Autosomal Dominant | CRANIOFACIAL MICROSOMIA 1 |
| SF3b130 | U12 snRNP | *SF3B3* | 16 | 70557691 | 70608820 | - | None |
| SF3b49 | U12 snRNP | *SF3B4* | 1 | 14989520<br>9 | 149900236 | Autosomal Dominant | ACROFACIAL DYSOSTOSIS 1, -GER TYPE |
| SF3b14a | U12 snRNP | *SF3B6* | 2 | 24290454 | 24299313 | - | None |
| SF3b14b | U12 snRNP | *PHF5A* | 22 | 41855721 | 41864729 | - | None |
| SF3b10 | U12 snRNP | *SF3B5* | 6 | 14441601<br>8 | 144416754 | - | None |
| U11/U12-31K | U12 snRNP | *ZCRB1* | 12 | 42705880 | 42719920 | - | None |
| U11/U12-65K | U12 snRNP | *RNPC3* | 1 | 10406831<br>3 | 104097861 | Autosomal Recessive | PITUITARY HORMONE DEFICIENCY, COMBINED, 7 |
| ZRSR2 | U12 snRNP | *ZRSR2* | X | 15808595 | 15841383 | - | None |
| CDC5L | nineteen complex (NTC) | *CDC5L* | 6 | 44355262 | 44418163 | - | None |
| SYF3 | nineteen complex (NTC) | *CRNKL1* | 20 | 20015012 | 20036690 | - | None |
| CWC15 | NTC-related (NTR) | *CWC15* | 11 | 94695787 | 94706776 | - | No Entry |
| SKIP | NTC-related (NTR) | *SNW1* | 14 | 78183942 | 78227550 | - | None |
| PLRG1 | NTC-related (NTR) | *PLRG1* | 4 | 15545615<br>8 | 155471587 | - | None |
| SNIP1 | retention and splicing (RES) complex | *SNIP1* | 1 | 38000050 | 38019903 | Autosomal Recessive | NEURODEVELOPMENTAL DISORDER WITH HYPOTONIA, CRANIOFACIAL ABNORMALITIES, AND SEIZURES |
| RBMX2 | retention and splicing (RES) complex | *RBMX2* | X | 12953594<br>3 | 129547317 | X-Linked Recessive | INTELLECTUAL DEVELOPMENTAL DISORDER, X-LINKED, SYNDROMIC, SHASHI TYPE (1 family) & INTELLECTUAL DEVELOPMENTAL DISORDER, X-LINKED, SYNDROMIC, GUSTAVSON TYPE (1 family) |
| BUD13 | retention and splicing (RES) complex | *BUD13* | 11 | 11661888<br>6 | 116643704 | - | None |
| PPIL2 | prolyl peptidyl isomerase (PPIase)–like proteins | *PPIL2* | 22 | 22006559 | 22054304 | - | None |
| CWC27 | prolyl peptidyl isomerase (PPIase)–like proteins | *CWC27* | 5 | 64064757 | 64314590 | Autosomal Recessive | RETINITIS PIGMENTOSA WITH OR WITHOUT SKELETAL ANOMALIES |

| | | | | | | | |
|---|---|---|---|---|---|---|---|
| PRP2 | Splicing Factors | *DHX16* | 6 | 30620896 | 30640814 | Autosomal Dominant | NEUROMUSCULAR OCULOAUDITORY SYNDROME |
| GPKOW | Splicing Factors | *GPKOW* | X | 48970334 | 48980151 | X-Linked Recessive | X-linked microhydranencephaly (unconfuirmed) |
| RNF113A | Splicing Factors | *RNF113A* | X | 119004497 | 119005791 | X-Linked Recessive | TRICHOTHIODYSTROPHY 5, NONPHOTOSENSITIVE |
| SRm300 | Splicing Factors | *SRRM2* | 16 | 2802330 | 2822539 | Autosomal Dominant | INTELLECTUAL DEVELOPMENTAL DISORDER, AUTOSOMAL DOMINANT 72 |
| CWC22 | Splicing Factors | *CWC22* | 2 | 180809603 | 180871840 | - | None |
| SRm160 | Splicing Factors | *SRRM1* | 1 | 24958207 | 24999758 | - | None |
| CRIPT | Splicing Factors | *CRIPT* | 2 | 46843555 | 46852881 | Autosomal Recessive | ROTHMUND-THOMSON SYNDROME, TYPE 3 |
| RBM48 | Splicing Factors | *RBM48* | 7 | 92158087 | 92167319 | - | No Entry |
| ARMC7 | Splicing Factors | *ARMC7* | 17 | 73106047 | 73126360 | - | No Entry |
| PRP3 | U4atac/U6atac di-snRNP | *PRPF3* | 1 | 150293925 | 150325671 | Autosomal Dominant | RETINITIS PIGMENTOSA 18 |
| PRP4 | U4atac/U6atac di-snRNP | *PRPF4* | 9 | 116037623 | 116055185 | Autosomal Dominant | RETINITIS PIGMENTOSA 70 |
| PRP31 | U4atac/U6atac di-snRNP | *PRPF31* | 19 | 54618837 | 54635140 | Autosomal Dominant | RETINITIS PIGMENTOSA 11 |
| SNU13 | U4atac/U6atac di-snRNP | *SNU13* | 22 | 42069934 | 42086508 | - | None |
| CE-TAC | U4atac/U6atac di-snRNP | *CE-TAC* | 11 | 118868852 | 118886501 | Autosomal Recessive | MOSAIC VARIEGATED ANEUPLOIDY SYNDROME 4 |
| LSM2 | U4atac/U6atac di-snRNP | *LSM2* | 6 | 31765173 | 31774761 | - | None |
| LSM3 | U4atac/U6atac di-snRNP | *LSM3* | 3 | 14219858 | 14242619 | - | None |
| LSM4 | U4atac/U6atac di-snRNP | *LSM4* | 19 | 18417040 | 18434084 | - | None |
| LSM5 | U4atac/U6atac di-snRNP | *LSM5* | 7 | 32524951 | 32534895 | - | None |
| LSM6 | U4atac/U6atac di-snRNP | *LSM6* | 4 | 147096837 | 147121152 | - | None |
| LSM7 | U4atac/U6atac di-snRNP | *LSM7* | 19 | 2321516 | 2328619 | - | None |
| LSM8 | U4atac/U6atac di-snRNP | *LSM8* | 7 | 117824086 | 117832878 | - | None |
| U11/U12-20K | U11 snRNP | *ZMAT5* | 22 | 30126945 | 30163000 | - | None |
| U11/U12-25K | U11 snRNP | *SNRNP25* | 16 | 103010 | 107669 | - | No Entry |
| U11/U12-35K | U11 snRNP | *SNRNP35* | 12 | 123942188 | 123957701 | - | None |
| U11/U12-48K | U11 snRNP | *SNRNP48* | 6 | 7590432 | 7612200 | - | No Entry |
| U11/U12-59K | U11 snRNP | *PDCD7* | 15 | 65409717 | 65426174 | - | None |
| SADI | Tri-snRNP specific | *USP39* | 2 | 85829979 | 85876403 | - | None |
| SNU66 | Tri-snRNP specific | *SART1* | 11 | 65729160 | 65747299 | - | None |
| PRP4 ki-se | Pre-B specific | *PRP4K* | 6 | 4021534 | 4065217 | - | None |

**Supplementary table 2:** Details of samples used for RNA-seq experiments. eT1D – early-onset type 1 diabetes. M – Male. F – Female.

| Sample | Condition | Sex | Age collected (years) | Age diagnosed with diabetes |
|---|---|---|---|---|
| T1D1 | eT1D control | M | 10-15 | 1-5 years |
| T1D2 | eT1D control | M | 6-10 | 0-6 months |
| T1D3 | eT1D control | F | 6-10 | 7-12 months |
| T1D4 | eT1D control | F | 6-10 | 1-5 years |
| HEAL1 | Healthy control | F | 1-5 | - |
| HEAL2 | Healthy control | M | 6-10 | - |
| HEAL3 | Healthy control | M | 1-5 | - |
| HEAL4 | Healthy control | F | 6-10 | - |
| 8 | *RNU4ATAC* affected | F | 6-10 | 0-6 months |
| 8 parent | *RNU4ATAC* carrier | M | 41-45 | - |
| 8 sibling | *RNU4ATAC* carrier | F | 16-20 | |
| Ci | *RNU6ATAC* affected | M | 1-5 | 0-6 months |
| C parent | *RNU6ATAC* carrier | F | 31-35 | - |
| C parent | *RNU6ATAC* carrier | M | 41-45 | - |
| Ciii | *RNU6ATAC* affected | F | 11-15 | 6-10 years |
| Cii | *RNU6ATAC* affected | M | 6-10 | 1-5 years |
| A parent | *RNU6ATAC* carrier | F | 26-30 | - |
| A parent | *RNU6ATAC* carrier | M | 25-30 | - |
| 11 | *RNU4ATAC* affected | F | 1-5 | 0-6 months |
| 11 parent | *RNU4ATAC* carrier | F | 26-30 | - |
| 11 parent | *RNU4ATAC* carrier | M | 36-40 | - |
| 6 | *RNU4ATAC* affected | F | 11-15 | 0-6 months |
| 6 parent | *RNU4ATAC* carrier | F | 46-50 | - |

**Supplementary table 3 –** U12-intron containing genes with intron retention determined from RNA-seq of patient whole blood erived RNA. Only genes with significant intron retention shown, which is defined by the magnitude of intron retention and the read depth over the transcript. IAOD – Intron Annotation and Orthology database (1). ND – not detected.

| **Gene** | **U12 intron reported in IAOD database** | **Intron retention in *RNU4ATAC* affected individuals** | **Mean normalised expression in *RNU4ATAC* affected** | **Intron retention in *RNU6ATAC* affected individuals** | **Mean normalised expression in *RNU6ATAC* affected** |
|---|---|---|---|---|---|
| *UPF1* | Yes | Yes | 5416 | Yes | 5253 |
| *RHBDD2* | Yes | Yes | 441 | Yes | 478 |
| *BAZ1B* | Yes | Yes | 3407 | Yes | 3640 |
| *CLCN6* | Yes | Yes | 778 | Yes | 728 |
| *PPP5C* | Yes | Yes | 702 | Yes | 648 |
| *CAPN1* | Yes | Yes | 2839 | Yes | 3098 |
| *CCDC28A* | Yes | Yes | 217 | Yes | 232 |
| *NUP160* | Yes | Yes | 1500 | Yes | 1332 |
| *ZCCHC8* | Yes | Yes | 1460 | Yes | 1490 |
| *EDC4* | Yes | Yes | 1161 | Yes | 1244 |
| *CYBA* | Yes | Yes | 5049 | Yes | 5645 |
| *CUL1* | Yes | Yes | 1083 | Yes | 1058 |
| *SBNO2* | Yes | Yes | 10655 | Yes | 10865 |
| *IP6K2* | Yes | Yes | 873 | Yes | 880 |
| *RASGRP2* | Yes | Yes | 4847 | Yes | 4599 |
| *PIGB* | Yes | Yes | 438 | Yes | 621 |
| *UFD1* | Yes | Yes | 1827 | Yes | 2248 |
| *WDR1* | Yes | Yes | 11352 | Yes | 12699 |
| *AP1M1* | Yes | Yes | 2142 | Yes | 2293 |
| *EED* | Yes | Yes | 582 | Yes | 581 |
| *SRI* | Yes | Yes | 654 | Yes | 691 |
| *KIFAP3* | Yes | Yes | 693 | Yes | 709 |
| *XRCC5* | Yes | Yes | 5101 | Yes | 4883 |
| *CCNT2* | Yes | Yes | 3550 | Yes | 3597 |
| *TNPO1* | Yes | Yes | 4441 | Yes | 6456 |
| *SMAP2* | Yes | Yes | 11222 | Yes | 10476 |
| *HAL* | Yes | Yes | 594 | Yes | 679 |
| *WDFY1* | Yes | Yes | 2860 | Yes | 1774 |
| *DDX18* | Yes | Yes | 1303 | Yes | 1335 |
| *SLC8B1* | Yes | Yes | 1444 | Yes | 1545 |
| *CCNK* | Yes | Yes | 1225 | Yes | 1456 |
| *MAEA* | Yes | Yes | 2448 | Yes | 2670 |
| *BTAF1* | Yes | Yes | 3082 | Yes | 3111 |

| | | | | | |
|---|---|---|---|---|---|
| *SRPK1* | Yes | Yes | 2378 | Yes | 2556 |
| *SETD1A* | Yes | Yes | 947 | Yes | 1013 |
| *HNRNPM* | Yes | Yes | 1981 | Yes | 2014 |
| *LZTR1* | Yes | Yes | 980 | Yes | 930 |
| *MAPK1* | Yes | Yes | 5603 | Yes | 6944 |
| *MYH9* | Yes | Yes | 45636 | Yes | 57808 |
| *PCNX1* | Yes | Yes | 5479 | Yes | 8193 |
| *CRNKL1* | Yes | Yes | 990 | Yes | 1086 |
| *SMS* | Yes | Yes | 467 | Yes | 526 |
| *VAC14* | Yes | Yes | 1001 | Yes | 935 |
| *CLCN7* | Yes | Yes | 2407 | Yes | 2150 |
| *TMEM87A* | Yes | Yes | 908 | Yes | 907 |
| *SPG11* | Yes | Yes | 5002 | Yes | 5188 |
| *INTS10* | Yes | Yes | 783 | Yes | 801 |
| *STX10* | Yes | Yes | 1078 | Yes | 1323 |
| *GARS1* | Yes | Yes | 1519 | Yes | 1491 |
| *RAPGEF1* | Yes | Yes | 6320 | Yes | 6878 |
| *HPS1* | Yes | Yes | 2202 | Yes | 2464 |
| *CUL2* | Yes | Yes | 1344 | Yes | 1232 |
| *GOSR1* | Yes | Yes | 1992 | Yes | 1817 |
| *NUP107* | Yes | Yes | 604 | Yes | 644 |
| *CHD4* | Yes | Yes | 5701 | Yes | 6314 |
| *SUDS3* | Yes | Yes | 1366 | Yes | 1595 |
| *MAPK14* | Yes | Yes | 6013 | Yes | 6475 |
| *MED23* | Yes | Yes | 2123 | Yes | 2079 |
| *SMAP1* | Yes | Yes | 919 | Yes | 1068 |
| *TMEM30A* | Yes | Yes | 2833 | Yes | 2955 |
| *IK* | Yes | Yes | 2283 | Yes | 2605 |
| *MSH3* | Yes | Yes | 433 | Yes | 423 |
| *NCBP2* | Yes | Yes | 919 | Yes | 913 |
| *ATP6V1A* | Yes | Yes | 3330 | Yes | 4383 |
| *SSR3* | Yes | Yes | 1432 | Yes | 1434 |
| *INO80D* | Yes | Yes | 5623 | Yes | 5727 |
| *WLS* | Yes | Yes | 290 | Yes | 414 |
| *RPF1* | Yes | Yes | 535 | Yes | 494 |
| *GBP1* | Yes | Yes | 10335 | Yes | 5377 |
| *STK11* | Yes | Yes | 1234 | Yes | 1603 |
| *TCP1* | Yes | Yes | 1218 | Yes | 1097 |
| *NAA60* | Yes | Yes | 1322 | Yes | 1392 |
| *THOC2* | Yes | Yes | 2413 | Yes | 2388 |
| *CAPNS1* | Yes | Yes | 2166 | Yes | 2465 |
| *HIP1* | Yes | Yes | 3104 | Yes | 3381 |
| *SPCS3* | Yes | Yes | 2968 | Yes | 3288 |
| *CCNT1* | Yes | Yes | 1803 | Yes | 2108 |
| *MAU2* | Yes | Yes | 2300 | Yes | 2449 |
| *FCHO1* | Yes | Yes | 851 | Yes | 946 |

| | | | | | |
|---|---|---|---|---|---|
| *ZC3H4* | Yes | Yes | 3031 | Yes | 3535 |
| *NOL11* | Yes | Yes | 889 | Yes | 854 |
| *DIAPH1* | Yes | Yes | 11991 | Yes | 14228 |
| *ACTR10* | Yes | Yes | 509 | Yes | 519 |
| *RAF1* | Yes | Yes | 15928 | Yes | 18837 |
| *NUP210* | Yes | Yes | 4653 | Yes | 4947 |
| *XPO4* | Yes | Yes | 1487 | Yes | 1463 |
| *ZFC3H1* | Yes | Yes | 4884 | Yes | 5219 |
| *RELCH* | Yes | Yes | 2164 | Yes | 2179 |
| *DOCK2* | Yes | Yes | 11074 | Yes | 11627 |
| *ERCC5* | Yes | Yes | 3286 | Yes | 3402 |
| *RINT1* | Yes | Yes | 405 | Yes | 426 |
| *SRPK2* | Yes | Yes | 3951 | Yes | 5518 |
| *DYNC1LI2* | Yes | Yes | 3651 | Yes | 3543 |
| *SMPD4* | Yes | Yes | 798 | Yes | 844 |
| *NCBP1* | Yes | Yes | 1216 | Yes | 1226 |
| *ARPC5L* | Yes | Yes | 638 | Yes | 562 |
| *DERL1* | Yes | Yes | 1685 | Yes | 1617 |
| *DCTN3* | Yes | Yes | 362 | Yes | 353 |
| *PLCB2* | Yes | Yes | 7071 | Yes | 8055 |
| *USO1* | Yes | Yes | 1708 | Yes | 1678 |
| *WDFY2* | Yes | Yes | 2253 | Yes | 2292 |
| *SBNO1* | Yes | Yes | 3017 | Yes | 3684 |
| *SETD1B* | Yes | Yes | 5810 | Yes | 6151 |
| *CUL4A* | Yes | Yes | 2198 | Yes | 2597 |
| *SLC12A6* | Yes | Yes | 8424 | Yes | 9498 |
| *PDPK1* | Yes | Yes | 2334 | Yes | 2209 |
| *RMC1* | Yes | Yes | 772 | Yes | 775 |
| *CNIH4* | Yes | Yes | 995 | Yes | 1021 |
| *PARP1* | Yes | Yes | 1942 | Yes | 1908 |
| *HNRNPLL* | Yes | Yes | 765 | Yes | 898 |
| *EML4* | Yes | Yes | 4880 | Yes | 5484 |
| *UBXN4* | Yes | Yes | 2679 | Yes | 2748 |
| *UBR3* | Yes | Yes | 2120 | Yes | 2116 |
| *DYNC1LI1* | Yes | Yes | 1326 | Yes | 1215 |
| *UBA3* | Yes | Yes | 954 | Yes | 1030 |
| *ATG3* | Yes | Yes | 2898 | Yes | 2173 |
| *MARCHF6* | Yes | Yes | 5966 | Yes | 7280 |
| *SLC30A5* | Yes | Yes | 807 | Yes | 791 |
| *SLC12A9* | Yes | Yes | 2591 | Yes | 2763 |
| *DIAPH2* | Yes | Yes | 1077 | Yes | 971 |
| *DOCK5* | Yes | Yes | 10466 | Yes | 13634 |
| *GOLGA7* | Yes | Yes | 1057 | Yes | 1213 |
| *FRA10AC1* | Yes | Yes | 505 | Yes | 588 |
| *INTS4* | Yes | Yes | 852 | Yes | 886 |
| *EML3* | Yes | Yes | 1172 | Yes | 1364 |

| | | | | | |
|---|---|---|---|---|---|
| *AHCTF1* | Yes | Yes | 5829 | Yes | 8171 |
| *TRAPPC8* | Yes | Yes | 1730 | Yes | 1837 |
| *GBP5* | Yes | Yes | 34638 | Yes | 14403 |
| *VBP1* | Yes | Yes | 354 | Yes | 394 |
| *ZDHHC5* | Yes | Yes | 2001 | Yes | 2017 |
| *MAPK13* | Yes | Yes | 1000 | Yes | 1346 |
| *FCHO2* | Yes | Yes | 2611 | Yes | 2941 |
| *BRAF* | Yes | Yes | 2579 | Yes | 3141 |
| *CUL4B* | Yes | Yes | 2412 | Yes | 2556 |
| *MORC3* | Yes | Yes | 7970 | Yes | 8154 |
| *SSU72* | Yes | Yes | 3187 | Yes | 3230 |
| *TRAPPC10* | Yes | Yes | 2838 | Yes | 2959 |
| *CRTC2* | Yes | Yes | 1343 | Yes | 1463 |
| *MYSM1* | Yes | Yes | 3959 | Yes | 4509 |
| *GBP2* | Yes | Yes | 12346 | Yes | 12902 |
| *CNST* | Yes | Yes | 3158 | Yes | 2940 |
| *CAPN2* | Yes | Yes | 3651 | Yes | 3625 |
| *RNF123* | Yes | Yes | 1277 | Yes | 1559 |
| *NAA15* | Yes | Yes | 1699 | Yes | 1588 |
| *ICE1* | Yes | Yes | 2790 | Yes | 2644 |
| *STXBP5* | Yes | Yes | 3526 | Yes | 5163 |
| *INTS8* | Yes | Yes | 1181 | Yes | 1324 |
| *TEPSIN* | Yes | Yes | 1191 | Yes | 1146 |
| *TRAPPC9* | Yes | Yes | 1095 | Yes | 1264 |
| *SRP68* | Yes | Yes | 712 | Yes | 771 |
| *C2CD3* | Yes | Yes | 1789 | Yes | 1799 |
| *STX8* | Yes | Yes | 260 | Yes | 301 |
| *HARS1* | Yes | Yes | 1166 | Yes | 1275 |
| *RASGRP4* | Yes | Yes | 1960 | Yes | 2290 |
| *RASGRP1* | Yes | Yes | 4824 | Yes | 4993 |
| *NAA16* | Yes | Yes | 1639 | Yes | 1559 |
| *PSME3IP1* | Yes | Yes | 1391 | Yes | 1627 |
| *DCP2* | Yes | Yes | 5769 | Yes | 6400 |
| *RALGAPA1* | Yes | Yes | 1744 | Yes | 1842 |
| *BRMS1* | Yes | Yes | 424 | Yes | 450 |
| *PPP2R2D* | Yes | Yes | 848 | Yes | 908 |
| *RAB6A* | Yes | Yes | 2469 | Yes | 2731 |
| *MLXIP* | Yes | Yes | 4204 | Yes | 3954 |
| *IP6K1* | Yes | Yes | 1530 | Yes | 1572 |
| *KCMF1* | Yes | Yes | 2112 | Yes | 2342 |
| *GAK* | Yes | Yes | 3012 | Yes | 3269 |
| *EIF3K* | Yes | Yes | 1206 | Yes | 1277 |
| *MED14* | Yes | Yes | 1289 | Yes | 1451 |
| *SLC9A9* | Yes | Yes | 641 | Yes | 581 |
| *RFX7* | Yes | Yes | 2351 | Yes | 2314 |
| *AP2A2* | Yes | Yes | 1366 | Yes | 1303 |

| | | | | | |
|---|---|---|---|---|---|
| *RNPC3* | Yes | Yes | 1178 | Yes | 1284 |
| *ZDHHC17* | Yes | Yes | 1901 | Yes | 1793 |
| *RNF220* | Yes | Yes | 1217 | Yes | 1276 |
| *TCEA1* | Yes | Yes | 1427 | Yes | 1693 |
| *RALGAPA2* | Yes | Yes | 4084 | Yes | 4991 |
| *AP2A1* | Yes | Yes | 2772 | Yes | 3968 |
| *MCMBP* | Yes | Yes | 980 | Yes | 1219 |
| *NOL8* | Yes | Yes | 1095 | Yes | 1140 |
| *UBL5* | Yes | Yes | 700 | Yes | 660 |
| *SLC9A6* | Yes | Yes | 727 | Yes | 828 |
| *ATG9A* | Yes | Yes | 1298 | Yes | 1487 |
| *INPP5B* | Yes | Yes | 980 | Yes | 830 |
| *ZDHHC18* | Yes | Yes | 3219 | Yes | 3777 |
| *CNEP1R1* | Yes | Yes | 623 | Yes | 701 |
| *SACM1L* | Yes | Yes | 3343 | Yes | 3335 |
| *ARL2* | Yes | Yes | 170 | Yes | 155 |
| *VPS16* | Yes | Yes | 471 | Yes | 554 |
| *PPP2R2A* | Yes | Yes | 1866 | Yes | 1581 |
| *VPS52* | Yes | Yes | 753 | Yes | 737 |
| *HSBP1* | Yes | Yes | 871 | Yes | 795 |
| *MCTS1* | Yes | Yes | 1061 | Yes | 1070 |
| *MYO1F* | No | Yes | 15962 | Yes | 18810 |
| *MTG1* | No | Yes | 447 | Yes | 394 |
| *CTNNBL1* | No | Yes | 546 | Yes | 521 |
| *GCA* | No | Yes | 7269 | Yes | 7902 |
| *SLC44A2* | No | Yes | 4857 | Yes | 6135 |
| *VPS9D1* | No | Yes | 2406 | Yes | 1832 |
| *ELOF1* | No | Yes | 533 | Yes | 533 |
| *POLR2E* | No | Yes | 1108 | Yes | 1275 |
| *PCID2* | No | Yes | 530 | Yes | 622 |
| *ARAF* | No | Yes | 926 | Yes | 1124 |
| *EXOSC9* | No | Yes | 396 | Yes | 418 |
| *PCNX3* | No | Yes | 2958 | Yes | 3067 |
| *INSIG1* | Yes | ND | 782 | Yes | 1139 |
| *NBPF26* | No | ND | 1716 | Yes | 1736 |
| *NBPF12* | No | ND | 792 | Yes | 844 |
| *GOLPH3L* | No | ND | 563 | Yes | 505 |
| *SCFD2* | No | ND | 314 | Yes | 285 |
| *PSMC4* | Yes | Yes | 510 | ND | 513 |
| *POLA2* | Yes | Yes | 461 | ND | 453 |
| *VEZT* | Yes | Yes | 666 | ND | 668 |
| *LSG1* | Yes | Yes | 1094 | ND | 839 |
| *TAF2* | Yes | Yes | 1090 | ND | 1189 |
| *IPO5* | Yes | Yes | 1399 | ND | 1335 |
| *VPS35* | Yes | Yes | 1735 | ND | 1668 |
| *DERL2* | Yes | Yes | 713 | ND | 684 |

| | | | | | |
|---|---|---|---|---|---|
| *EIF4G3* | Yes | Yes | 2776 | ND | 2386 |
| *RABL2B* | Yes | Yes | 595 | ND | 574 |
| *EIF3I* | Yes | Yes | 844 | ND | 864 |
| *ASCC2* | Yes | Yes | 2634 | ND | 4654 |
| *FKBP3* | Yes | Yes | 327 | ND | 349 |
| *USP14* | Yes | Yes | 815 | ND | 1004 |
| *MAGT1* | Yes | Yes | 1024 | ND | 1109 |
| *TNPO2* | Yes | Yes | 658 | ND | 743 |
| *CRTC1* | Yes | Yes | 567 | ND | 509 |
| *LSM5* | Yes | Yes | 164 | ND | 144 |
| *SMC3* | Yes | Yes | 1964 | ND | 1946 |
| *DRG2* | Yes | Yes | 451 | ND | 467 |
| *TMEM33* | Yes | Yes | 1859 | ND | 2073 |
| *GAR1* | Yes | Yes | 124 | ND | 134 |
| *E2F3* | Yes | Yes | 1271 | ND | 1348 |
| *FIG4* | Yes | Yes | 1728 | ND | 1392 |
| *HARS2* | Yes | Yes | 614 | ND | 667 |
| *SLC12A7* | Yes | Yes | 1329 | ND | 963 |
| *SPCS2* | Yes | Yes | 589 | ND | 563 |
| *WDR11* | Yes | Yes | 1317 | ND | 1353 |
| *EML2* | Yes | Yes | 348 | ND | 374 |
| *LSM8* | Yes | Yes | 1690 | ND | 1772 |
| *PSMA1* | Yes | Yes | 1101 | ND | 1021 |
| *XPO7* | Yes | Yes | 3127 | ND | 3236 |
| *ATXN10* | Yes | Yes | 559 | ND | 549 |
| *EXOSC2* | Yes | Yes | 723 | ND | 666 |
| *PPIL4* | Yes | Yes | 689 | ND | 659 |
| *VPS25* | Yes | Yes | 206 | ND | 235 |
| *DAP3* | Yes | Yes | 620 | ND | 722 |
| *NAT10* | Yes | Yes | 717 | ND | 764 |
| *STX6* | Yes | Yes | 1141 | ND | 1138 |
| *KANSL2* | Yes | Yes | 549 | ND | 535 |
| *USP21* | Yes | Yes | 431 | ND | 449 |
| *RFX5* | Yes | Yes | 1139 | ND | 963 |
| *RABL2A* | Yes | Yes | 422 | ND | 400 |
| *TMEM87B* | Yes | Yes | 631 | ND | 697 |
| *NUP205* | Yes | Yes | 1462 | ND | 1186 |
| *LSM12* | Yes | Yes | 680 | ND | 799 |
| *GBP4* | Yes | Yes | 6942 | ND | 3765 |
| *SLC66A3* | Yes | Yes | 228 | ND | 286 |
| *SSR2* | Yes | Yes | 1400 | ND | 1677 |
| *NEPRO* | Yes | Yes | 1091 | ND | 1071 |
| *MIOS* | Yes | Yes | 464 | ND | 479 |
| *SPTSSA* | Yes | Yes | 194 | ND | 283 |
| *TMEM41B* | Yes | Yes | 701 | ND | 623 |
| *DDB1* | Yes | Yes | 2655 | ND | 2754 |

| | | | | | |
|---|---|---|---|---|---|
| *CHD3* | Yes | Yes | 4662 | ND | 5875 |
| *EXOSC1* | Yes | Yes | 301 | ND | 344 |
| *NAA20* | Yes | Yes | 240 | ND | 231 |
| *SRP72* | Yes | Yes | 1483 | ND | 1526 |
| *DRAP1* | Yes | Yes | 605 | ND | 529 |
| *TSPYL2* | Yes | Yes | 653 | ND | 544 |
| *POLR3C* | Yes | Yes | 705 | ND | 694 |
| *GPR89B* | Yes | Yes | 692 | ND | 778 |
| *SRSF10* | Yes | Yes | 3592 | ND | 3386 |
| *HTT* | Yes | Yes | 3282 | ND | 3369 |
| *E2F4* | Yes | Yes | 1223 | ND | 1268 |
| *RECQL5* | No | Yes | 506 | ND | 477 |

**Supplementary table 4:** samples and controls for methylation analysis

| ID | Status | Age collected |
|---|---|---|
| 1 | *RNU4ATAC* | 0-6 months |
| 2 | *RNU4ATAC* | 1-5 years |
| 3 | *RNU4ATAC* | 7-12 months |
| 4 | *RNU4ATAC* | 0-6 months |
| 5 | *RNU4ATAC* | 7-12 months |
| 6 | *RNU4ATAC* | 0-6 months |
| 7 | *RNU4ATAC* | 1-5 years |
| 8 | *RNU4ATAC* | 6-10 years |
| 9 | *RNU4ATAC* | 7-12 months |
| 12 | *RNU4ATAC* | 0-6 months |
| Aii | *RNU6ATAC* | 0-6 months |
| Aii | *RNU6ATAC* | 21-25 years |
| B | *RNU6ATAC* | 0-6 months |
| Ci | *RNU6ATAC* | 0-6 months |
| Cii | *RNU6ATAC* | 11-15 years |
| Ciii | *RNU6ATAC* | 6-10 years |
| D | *RNU6ATAC* | 1-5 years |
| H1 | Healthy Control | 0-6 months |
| H10 | Healthy Control | 1-5 years |
| H11 | Healthy Control | 1-5 years |
| H12 | Healthy Control | 1-5 years |
| H13 | Healthy Control | 6-10 years |
| H14 | Healthy Control | 11-15 years |
| H15 | Healthy Control | 11-15 years |
| H16 | Healthy Control | 16-20 years |
| H17 | Healthy Control | 21-25 years |
| H2 | Healthy Control | 0-6 months |
| H3 | Healthy Control | 0-6 months |
| H4 | Healthy Control | 0-6 months |
| H5 | Healthy Control | 7-12 months |
| H6 | Healthy Control | 7-12 months |
| H7 | Healthy Control | 7-12 months |
| H8 | Healthy Control | 1-5 years |
| H9 | Healthy Control | 1-5 years |

**Supplementary Table 5:** Monoclonal antibodies used in the flow cytometry panels

| Marker (surface) | Fluorochrome | Clone | Company | Panel |
|---|---|---|---|---|
| Brilliant stain buffer | N/A | N/A | BD Biosciences | All |
| CD3 | APCCy7 | OKT3 | BioLegend | T cell |
| CD4 | BUV395 | SK3 | BD Biosciences | T cell |
| CD25 | BB515 | 2A3 | BD Biosciences | T cell |
| CD25 | BB515 | M-A251 | BD Biosciences | T cell |
| CD127 | APC | A019D5 | BioLegend | T cell |
| CD45RA | BV785 | HI100 | BioLegend | T cell |
| CD197/CCR7 | BV421 | G043H7 | BioLegend | T cell |
| CD183/CXCR3 | BV510 | G025H7 | BioLegend | T cell |
| CD194/CCR4 | BV605 | L291H4 | BioLegend | T cell |
| CD196/CCR6 | BUV737 | 11A9 | BD Biosciences | T cell |
| CD95 | PE | DX2 | BioLegend | T cell |
| CCR10 | PerCP-Cy5.5 | 1B5 | BD Biosciences | T cell |
| CD185/CXCR5 | PE-Cy7 | J252D4 | BioLegend | T cell |
| CD278/ICOS | BV711 | DX29 | BD Biosciences | T cell |
| CD279/PD-1 | PE-Dazzle 594 | EH12.2H7 | BioLegend | T cell |
| CD3 | APC-Cy7 | OKT3 | BioLegend | B/DC/Monocyte |
| CD19 | PE | HIB19 | BioLegend | B/DC/Monocyte |
| CD14 | PerCP-Cy5.5 | HCD14 | BioLegend | B/DC/Monocyte |
| CD16 | PE-Cy7 | 3G8 | BioLegend | B/DC/Monocyte |
| CD56 | APC | HCD56 | BioLegend | B/DC/Monocyte |
| HLA-DR | FITC | LN3 | BioLegend | B/DC/Monocyte |
| CD123 | BUV395 | 7G3 | BD Biosciences | B/DC/Monocyte |
| CD11c | BV421 | Bu15 | BioLegend | B/DC/Monocyte |
| IgD | BV605 | IA6-2 | BioLegend | B/DC/Monocyte |
| CD27 | BUV737 | L128 | BD Biosciences | B/DC/Monocyte |
| CD38 | BV785 | HIT2 | BioLegend | B/DC/Monocyte |
| CD24 | BV510 | ML5 | BioLegend | B/DC/Monocyte |
| CD45 | PerCP | HI30 | BioLegend | Granulocyte |
| CD3 | APC-Cy7 | OKT3 | BioLegend | Granulocyte |
| CD19 | APC-Cy7 | HIB19 | BioLegend | Granulocyte |
| CD56 | APC-Cy7 | HCD56 | BioLegend | Granulocyte |
| CD14 | AF488 | HCD14 | BioLegend | Granulocyte |
| CD15 | BV605 | W6D3 | BioLegend | Granulocyte |
| CD64 | BV421 | 10.1 | BioLegend | Granulocyte |
| CD63 | APC | HC56 | BioLegend | Granulocyte |
| CD123 | BUV395 | 7G3 | BD Biosciences | Granulocyte |
| CD294 | PE | BM16 | Biolegend | Granulocyte |
| CD203c | BV510 | NP4D6 | BioLegend | Granulocyte |
| CD69 | PE-Cy7 | FN50 | BioLegend | Granulocyte |
| CD45 | PerCP | HI30 | BioLegend | Lineage |
| CD3 | APC-Cy7 | OKT3 | BioLegend | Lineage |

| | | | | |
|---|---|---|---|---|
| CD4 | BUV395 | SK3 | BD Biosciences | Lineage |
| CD8 | BUV737 | SK1 | BD Biosciences | Lineage |
| CD19 | PE | HIB19 | BioLegend | Lineage |
| CD14 | AF488 | HCD14 | BioLegend | Lineage |
| CD16 | PE-Cy7 | 3G8 | BioLegend | Lineage |
| CD15 | BV605 | W6D3 | BioLegend | Lineage |
| CD56 | APC | HCD56 | BioLegend | Lineage |
| CD45RA | BV785 | HI100 | BioLegend | Treg |
| CD15s | BV510 | CSLEX1 | BD Biosciences | Treg |
| CD3 | BV605 | OKT3 | BioLegend | Treg |
| CD4 | BUV395 | SK3 | BD Biosciences | Treg |
| CD8 | BUV737 | SK1 | BD Biosciences | Treg |
| CD25 | PE | M-A251 | BD Biosciences | Treg |
| FOXP3 | AF647 | 259D | Beckman Coulter | Treg |
| Helios | Pacific Blue | 22F6 | BioLegend | Treg |
| Ki67 | FITC | B56 | BD Biosciences | Treg |
| CD69 | PE-Cy7 | FN50 | BioLegend | Treg |

**Supplementary table 6:** Markers used to define cell populations from flow cytometry

| | |
|---|---|
| Total B cells | CD45+CD15-CD3-CD19+ |
| IgD+ B cells | CD3-CD19+IgD+ |
| Immature B cells | CD3-CD19+CD27-CD24hiCD38hi |
| Mature Naïve B cells | CD3-CD19+CD27-CD24+CD38h+ |
| Memory B cells | CD3-CD19+CD27+CD24+CD38lo/- |
| Plasmablasts/Antibody secreting cells (ASCs) | CD3-CD19+CD27+CD24-CD38+/hi |
| Natural Killer (NK) cells | CD3CD19-CD14-CD56+ |
| Total Dendritic Cells (DC) | CD3-CD19-CD14-CD16-CD56lo/-HLA-DR+ |
| Myeloid DC | CD3-CD19-CD14-CD16-CD56lo/-HLA-DR+CD11c+CD123- |
| Plasmacytoid DC | CD3-CD19-CD14-CD16-CD56lo/-HLA-DR+CD11-CD123+ |
| Intermediate monocytes | CD3-CD19-CD56-/loHLA-DR+CD14+CD16+ |
| Classical monocytes | CD3-CD19-CD56-/loHLA-DR+CD14+CD16- |
| Non classical monocytes | CD3-CD19-CD56-/loHLA-DR+CD14lo/-CD16+ |
| FOXP3+ Tregs | CD3+CD4+CD25+FOXP3+ |
| Activated regulatory T cell (aTreg) | CD3+CD4+CD25+FOXP3hiCD45RAlo/- |
| Memory Treg (mTreg) | CD3+CD4+CD25+FOXP3+CD45RAlo/- |
| Resting Treg (rTreg) | CD3+CD4+CD25+FOXP3+ CD45RA+ |
| CD8 T cells | CD45+CD15-CD19-CD56-CD3+CD4-CD8+ |
| CD4 T cells | CD45+CD15-CD19-CD56-CD3+CD+CD8- |
| Conventional T cell (Tconv) | CD45+CD15-CD19-CD56-CD3+ |
| Central memory T cell (Tcm) | CD3+CD4+CD127+/hiCD25lo/-CCR7+CD45RA- |
| Terminally differentiated effector memory T cell (Temra) | CD3+CD4+CD127+/hiCD25lo/-CCR7-CD45RA+ |
| T effector memory (Tem) | CD3+CD4+CD127+/hiCD25lo/-CCR7-CD45RA- |
| Naïve T cell (Tn) | CD3+CD4+CD127+/hiCD25lo/-CD95-CCR7+CD45RAhi |
| T helper cell type 1 (Th1) | CD3+CD4+CD127+/hiCD25lo/-CCR7varCD45RAvarCXCR5-CCR4-CXCR3+CCR10-CCR6- |
| T helper cell type 1+ (Th1+) | CD3+CD4+CD127+/hiCD25lo/-CCR7varCD45RAvarCXCR5-CCR4-CXCR3+CCR10-CCR6+ |
| T helper cell type 2 (Th2) | CD3+CD4+CD127+/hiCD25lo/-CCR7varCD45RAvarCXCR5-CCR4+CXCR3-CCR10-CCR6- |
| T helper cell type 17 (Th17) | CD3+CD4+CD127+/hiCD25lo/-CCR7varCD45RAvarCXCR5-CCR4+CXCR3-CCR10-CCR6+ |
| T helper cell type 22 (Th22) | CD3+CD4+CD127+/hiCD25lo/-CCR7varCD45RAvarCXCR5-CCR4+CXCR3-CCR10+CCR6+ |
| Total Tregs | CD3+CD4+CD127-CD25+ |
| Treg central memory (cm) | CD3+CD4+CD127-CD25+CCR7+CD45RA- |
| Treg Temra | CD3+CD4+CD127-CD25+CCR7-CD45RA+ |
| Treg effector memory (em) | CD3+CD4+CD127-CD25+CCR7-CD45R- |
| Naïve Tregs | CD3+CD4+CD127-CD25+CD95-CCR7+CD45RAhi |
| Treg Th1 | CD3+CD4+CD127-CD25l+CCR7varCD45RAvarCXCR5-CCR4-CXCR3+CCR10-CCR6- |
| Treg Th1+ | CD3+CD4+CD127-CD25l+CCR7varCD45RAvarCXCR5-CCR4-CXCR3+CCR10-CCR6+ |
| Treg Th2 | CD3+CD4+CD127-CD25l+CCR7varCD45RAvarCXCR5-CCR4+CXCR3-CCR10-CCR6- |

| | |
|---|---|
| Treg Th17 | CD3+CD4+CD127-CD25l+CCR7varCD45RAvarCXCR5-CCR4+CXCR3-CCR10-CCR6+ |
| Treg Th22 | CD3+CD4+CD127-CD25l+CCR7varCD45RAvarCXCR5-CCR4+CXCR3-CCR10+CCR6+ |
| Eosinophils | CD45+CD3-CD14-CD19-CD56-CD15+CD294+CD203c+ |
| Neutrophils | CD45+CD3-CD14-CD19-CD56-CD15+CD294-CD203clo/- |
| Basophils | CD45+CD3-CD14-CD19-CD56-CD15-CD123+CD294+ |
| CD69+Tregs | CD3+CD4+CD25+FOXP3+CD69+ |
| CD69+ effector T cells (Teff) | CD3+CD4+CD25-FOXP3-CD69+ |
| CD69+ CD8 T cells | CD3+CD-CD8+CD69+ |
| Proliferating Tregs | CD3+CD4+CD25+FOXP3+Ki67+ |
| Proliferating CD4 Teff | CD3+CD4+CD25-FOXP3- Ki67+ |
| Proliferating CD8 T cells | CD3+CD4+CD25-FOXP3- Ki67+ |

**Supplementary table 7:** controls for flow cytometry

| ID | Type | Age collected | Age diagnosed diabetes |
|---|---|---|---|
| HC1 | Healthy | 11-15 years | - |
| HC2 | Healthy | 6-10 years | - |
| HC3 | Healthy | 6-10 years | - |
| HC4 | Healthy | 11-15 years | - |
| T1D1 | Type 1 diabetes | 11-15 years | 11-15 years |
| T1D2 | Type 1 diabetes | 11-15 years | 11-15 years |
| T1D3 | Type 1 diabetes | 11-15 years | 11-15 years |
| T1D4 | Type 1 diabetes | 11-15 years | 11-15 years |
| T1D5 | Type 1 diabetes | 11-15 years | 11-15 years |

**Supplementary figure 1:** Coverage plots showing genes with significant intron retention in whole blood RNA from individuals with biallelic pathogenic variants in *RNU4ATAC*, *RNU6ATAC* or both (supplementary table 3) but not present in the IAOD database (https://introndb.lerner.ccf.org/) (1).

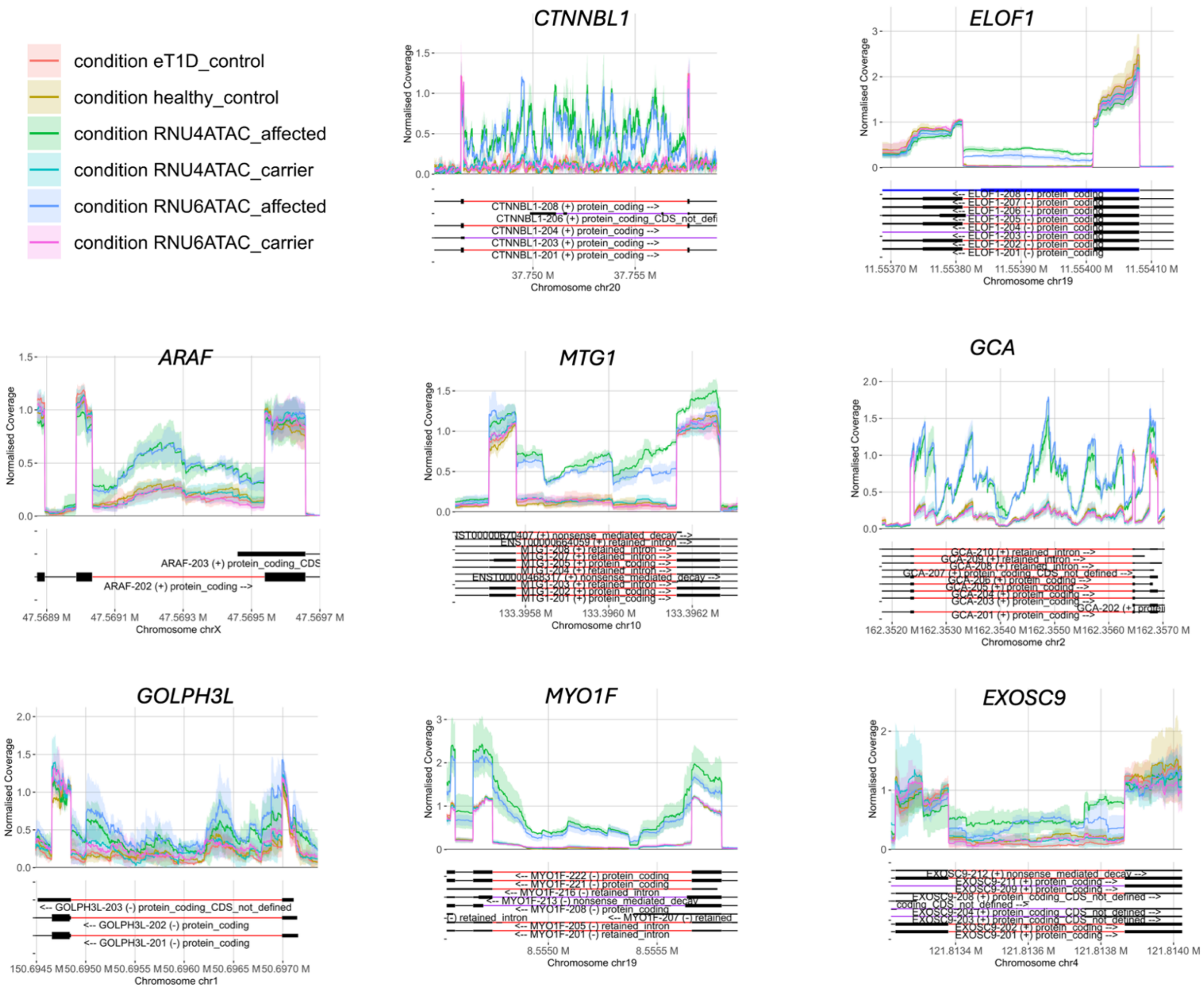

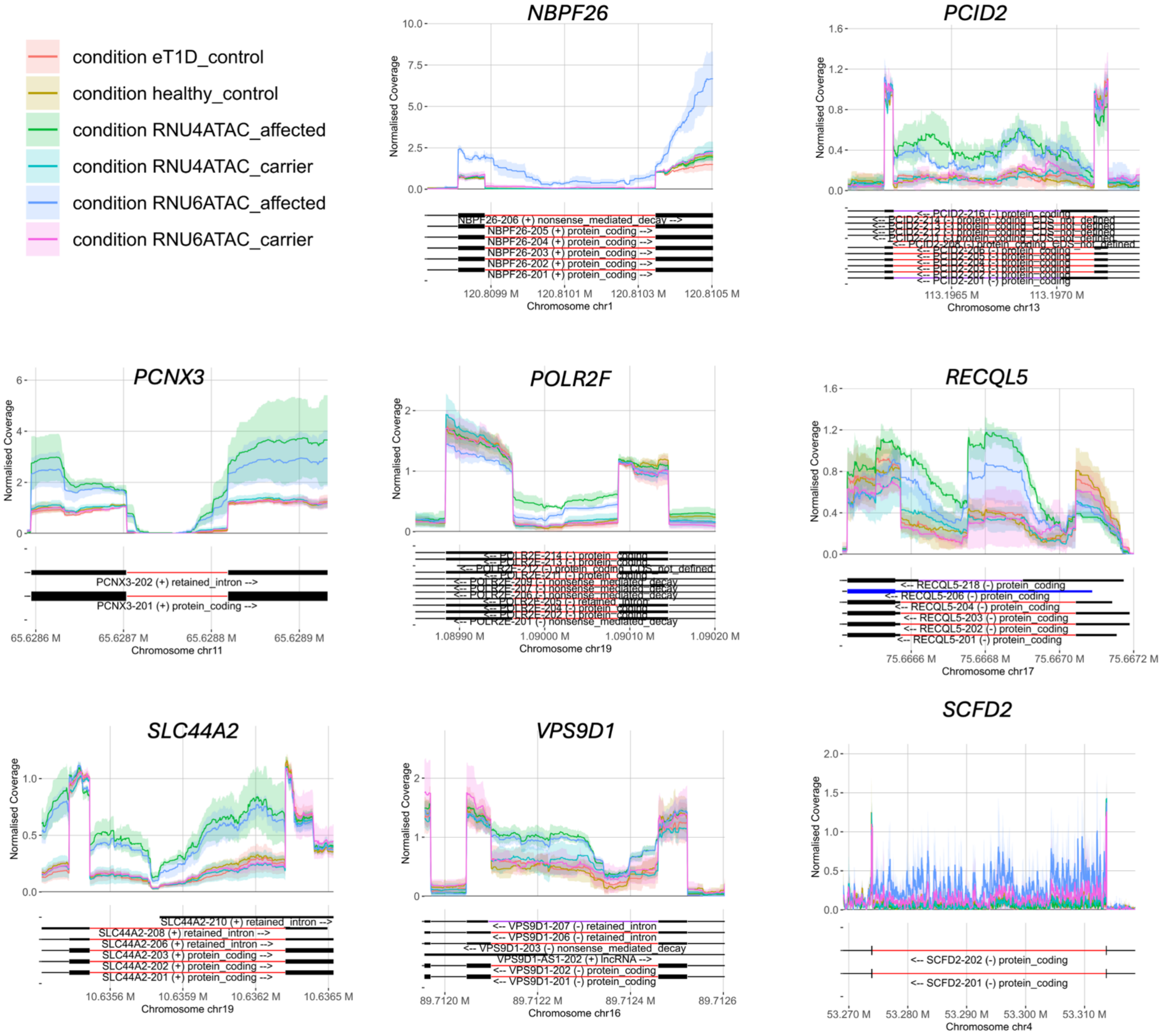
condition eT1D_control
condition healthy_control
condition RNU4ATAC_affected
condition RNU4ATAC_carrier
condition RNU6ATAC_affected
condition RNU6ATAC_carrier
NBPF26
Normalised Coverage
NBPF26-206 (+) nonsense_mediated_decay -->
NBPF26-205 (+) protein_coding -->
NBPF26-204 (+) protein_coding -->
NBPF26-203 (+) protein_coding -->
NBPF26-202 (+) protein_coding -->
NBPF26-201 (+) protein_coding -->
120.8099 M
120.8101 M
120.8103 M
120.8105 M
Chromosome chr1
PCID2
<-- PCID2-216 (-) protein_coding
<-- PCID2-201 (-) protein_coding
113.1965 M
113.1970 M
Chromosome chr13
PCNX3
PCNX3-202 (+) retained_intron -->
PCNX3-201 (+) protein_coding -->
65.6286 M
65.6287 M
65.6288 M
65.6289 M
Chromosome chr11
POLR2F
<-- POLR2F-212 (-) protein_coding_CDS_not_defined
<-- POLR2F-205 (-) retained_intron
<-- POLR2F-201 (-) nonsense_mediated_decay
1.08990 M
1.09000 M
1.09010 M
1.09020 M
Chromosome chr19
RECQL5
<-- RECQL5-218 (-) protein_coding
<-- RECQL5-206 (-) protein_coding
<-- RECQL5-204 (-) protein_coding
<-- RECQL5-203 (-) protein_coding
<-- RECQL5-202 (-) protein_coding
<-- RECQL5-201 (-) protein_coding
75.6666 M
75.6668 M
75.6670 M
75.6672 M
Chromosome chr17
SLC44A2
SLC44A2-210 (+) retained_intron -->
SLC44A2-208 (+) retained_intron -->
SLC44A2-206 (+) retained_intron -->
SLC44A2-203 (+) protein_coding -->
SLC44A2-202 (+) protein_coding -->
SLC44A2-201 (+) protein_coding -->
10.6356 M
10.6359 M
10.6362 M
10.6365 M
Chromosome chr19
VPS9D1
<-- VPS9D1-207 (-) retained_intron
<-- VPS9D1-206 (-) retained_intron
<-- VPS9D1-203 (-) nonsense_mediated_decay
VPS9D1-AS1-202 (+) lncRNA -->
<-- VPS9D1-202 (-) protein_coding
<-- VPS9D1-201 (-) protein_coding
89.7120 M
89.7122 M
89.7124 M
89.7126
Chromosome chr16
SCFD2
<-- SCFD2-202 (-) protein_coding
<-- SCFD2-201 (-) protein_coding
53.270 M
53.280 M
53.290 M
53.300 M
53.310 M
Chromosome chr4

**Supplementary figure 2**: Enrichment analysis of WGCNA gene modules. For the two significant modules, GO and KEGG enrichment was used to identify pathways connected to the gene lists.

A)

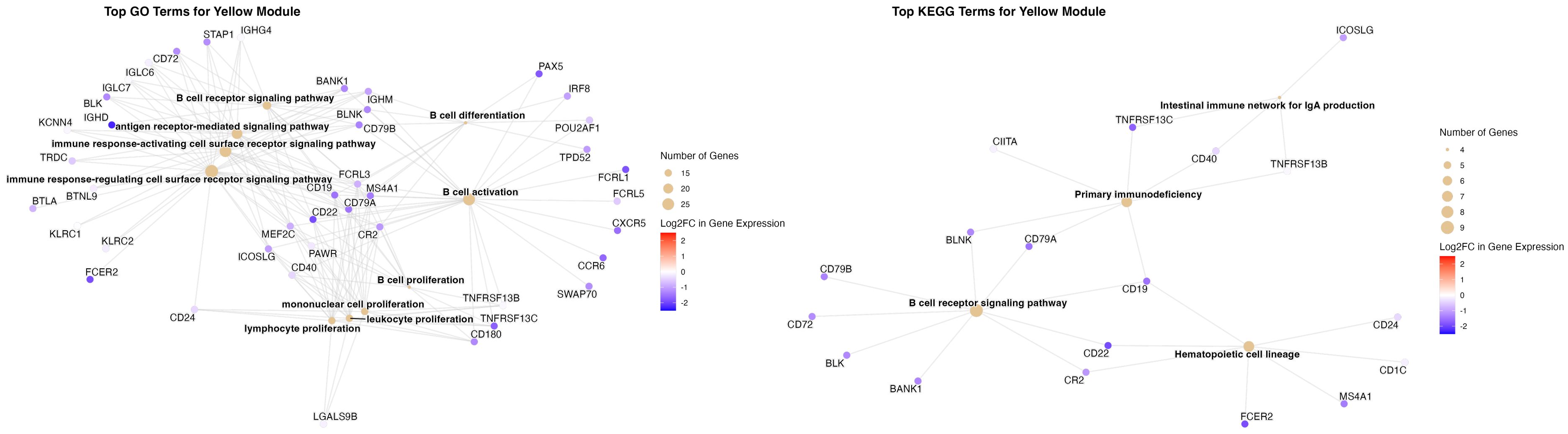


B)

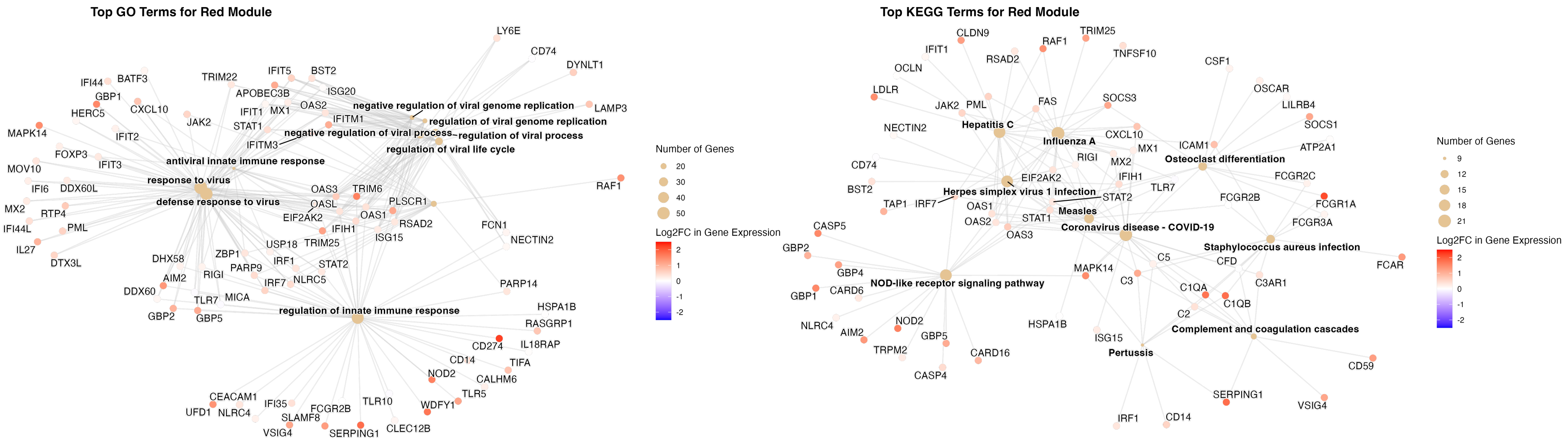

**Supplementary figure 3** Estimates of immune cell proportions from deconvolution of EPIC array methylation analysis of whole blood-derived DNA.

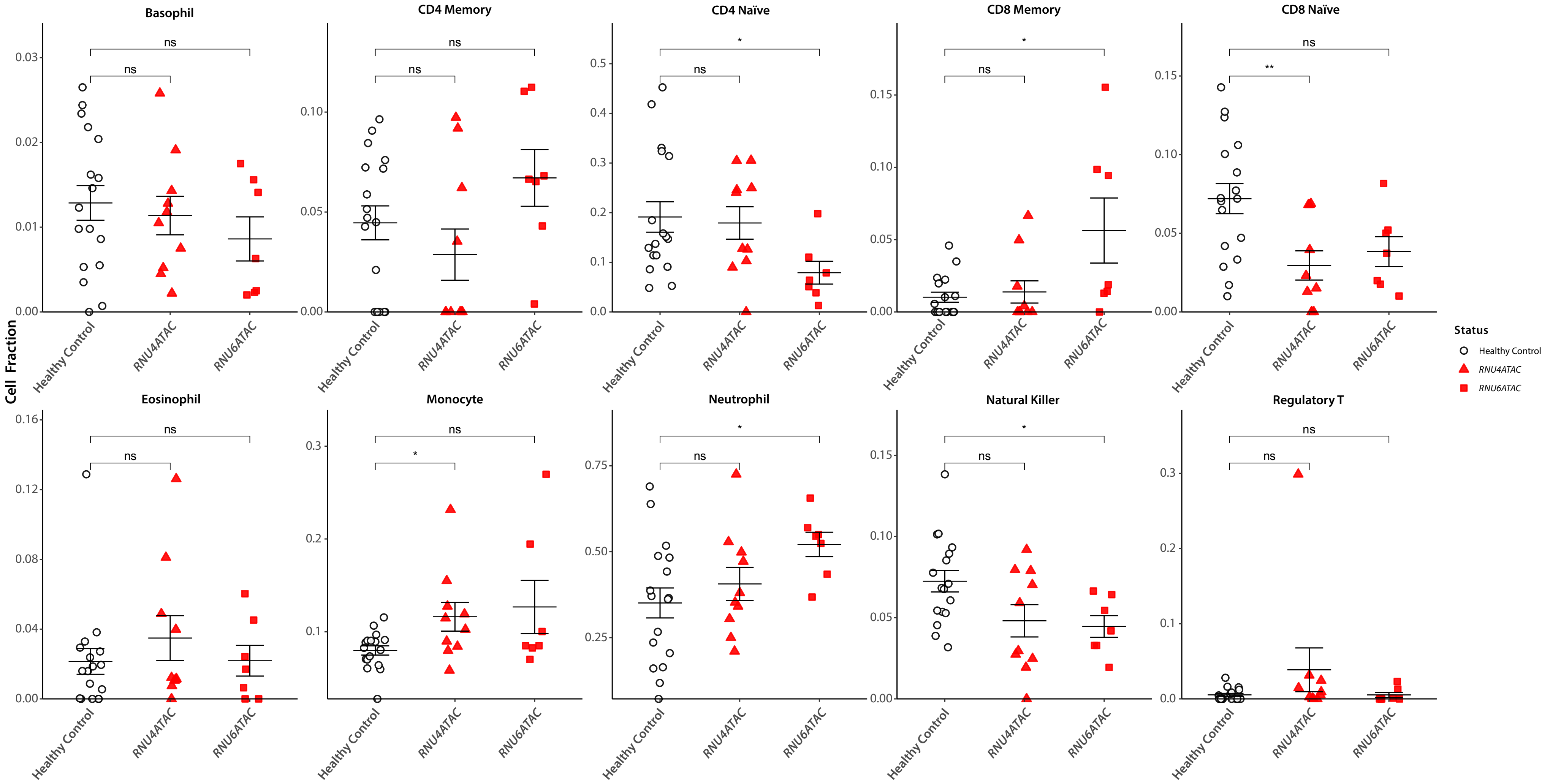

**Supplementary figure 4**: Immune phenotyping of *RNU4ATAC* case (patient 6), their unaffected heterozygous carrier parent, and age matched controls.

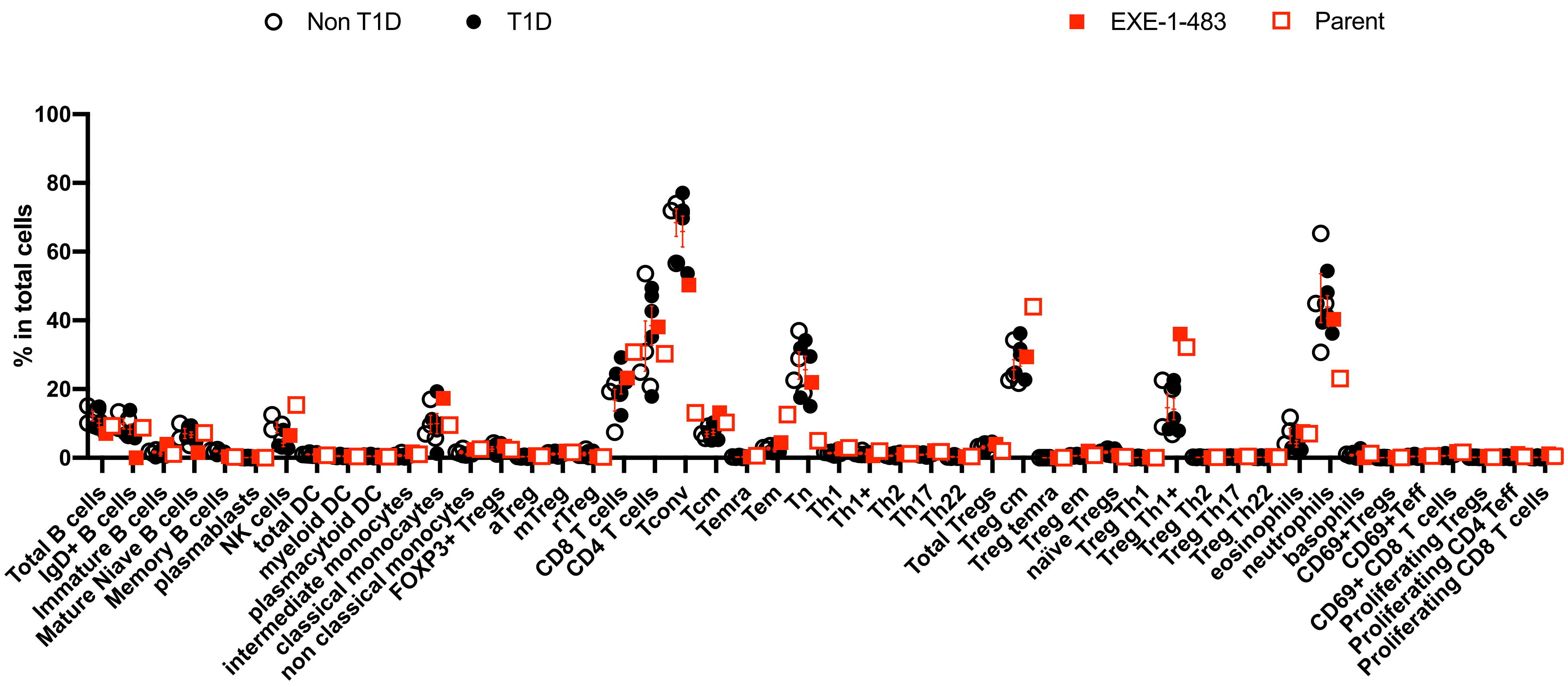
